# A retrospective study of a Chinese vision-language large model for emergency 3D brain CT interpretation

**DOI:** 10.64898/2026.07.11.26357421

**Authors:** Yifei Chen, Jingyuan Zheng, Yuanhan Wang, Beining Wu, Lu Li, Mingxuan Liu, Liaoman Xu, Yueyi Wu, Chang Liu, Lulu Guo, Hongjia Yang, Xuguang Bai, Feiwei Qin, Qiang Liao, Yong Gu, Gang Zhao, Lu Ma, Keqin Pan, Jun Guo, Ying Zhou, Huaiqiang Sun, Qiyuan Tian

**Affiliations:** Department of Radiology, Institution of Radiology and Medical Imaging, West China Hospital of Sichuan University, Chengdu, 610041, China; School of Biomedical Engineering, Tsinghua University, Beijing, 100084, China; School of Computer Science and Technology, Hangzhou Dianzi University, Hangzhou, 310018, China; Beichuan Qiang Autonomous County People’ s Hospital, Mianyang, 622750, China; Santai County People’ s Hospital, Mianyang, 621100, China; Tsinghua University Hospital, Beijing, 100084, China; Department of Radiology, Mianyang Central Hospital, Mianyang, 621000, China

## Abstract

Emergency brain computed tomography (CT) is the first line imaging modality for patients with acute neurological symptoms and trauma, where delayed or incomplete recognition of critical findings can directly compromise clinical outcomes. However, emergency CT interpretation and Chinese reporting remain highly variable under severe time constraints and heterogeneous institutional settings. In this study, we develop ERBrain, a multimodal large model specifically tailored for emergency brain CT, which jointly performs three-dimensional image understanding, Chinese radiology report generation, and emergency severity triage within a unified framework. ERBrain integrates volumetric visual representations with a Chinese large language model and explicitly prioritizes emergency critical signs through risk focused training objectives and a lightweight knowledge-augmented prompting strategy. Using more than 10,000 multicentre emergency CT studies, ERBrain achieved an accuracy of 0.943 and a balanced accuracy of 0.940 for three-level emergency triage and achieved the highest FIES-Avg clinical semantic score among the evaluated report-generation models in the in-distribution cohort. Across external data, ERBrain maintained favourable triage performance in two cross-institutional validation cohorts, whereas performance was lower but remained clinically informative in a third cohort characterized by an extremely low prevalence of Positive cases. These findings support further prospective evaluation of ERBrain as a radiology worklist prioritization and report-drafting assistant in heterogeneous emergency imaging settings.

## Introduction

Emergency brain computed tomography (CT) is the first-line examination for acute neurological conditions, where rapid and accurate interpretation drives life-saving decisions under severe time pressure^1–4^. Clinicians must identify life-threatening signs, such as subarachnoid hemorrhage, epidural or subdural hematoma, midline shift, and brain herniation, from three-dimensional volumetric data within minutes, and then communicate these findings clearly to emergency decision-making teams through reports^5–7^. This process relies heavily on individual expertise and sustained attention, making it vulnerable to missed findings, delayed diagnoses, and inconsistent report quality. These risks intensify under real-world constraints, such as specialist shortages in primary hospitals and rising emergency workloads in tertiary centers^8^. Therefore, improving the consistency, speed, and clinical usability of emergency brain CT interpretation without increasing physicians’ workload has become critical for building resilient emergency care systems^9,10^.

In recent years, multimodal large models have driven rapid progress in medical image interpretation, particularly in routine and relatively standardized clinical settings^11^. Trained on large-scale image–text pairs, these models couple visual understanding with clinically relevant language generation. This capability enables a wide range of radiology applications^12,13^, including abnormality detection, lesion segmentation, report generation, and clinical decision support across modalities, such as chest X-ray and abdominal CT^14–17^. These advances highlight the potential of a unified multimodal framework to bridge image analysis with downstream clinical communication and reasoning. They also suggest that such models could provide a promising starting point for brain CT interpretation, and potentially for emergency brain CT, where imaging findings must be translated into timely and clinically actionable decisions^18^. Nevertheless, whether current multimodal large models can fully meet the demands of emergency neuroimaging remains uncertain, given the need for comprehensive volumetric brain understanding and precise characterization of acute critical findings^19^.

Emergency neuroimaging requires multimodal large models to move beyond slice-level understanding toward full volumetric brain reasoning that directly supports time-critical clinical decision-making. Most existing multimodal large models use two-dimensional images or single-slice CT as input. Only a limited number of models incorporate volumetric data, but they are typically developed for general whole-body imaging focused on common conditions and chronic disease trajectories^20,21^. As a result, they fail to capture the spatiotemporal distribution of high-risk acute findings, such as active hemorrhage, evolving cerebral edema, and intracranial pressure crises, across the full brain volume^22,23^. This insufficient volumetric modeling of acute critical findings fundamentally limits downstream brain CT report generation, confining models to coarse assessments of abnormality presence or broad diagnostic impressions. They also cannot explicitly model emergency priority, potential intervention needs, or the coexistence of multiple lesions^24,25^. For emergency physicians, omission of time-critical information, such as whether midline shift exceeds five millimeters or whether intracranial hemorrhage is present, can compromise time-sensitive management even when the overall impression appears correct^26–28^. Consequently, relying solely on general-purpose architectures and conventional task formulations is unlikely to meet the combined demands of high sensitivity, high specificity, and strong clinical actionability required in emergency neuroimaging^29^.

Moreover, medical report generation is still predominantly evaluated using general-purpose language metrics that fail to capture whether radiology reports accurately convey critical findings, diagnostic conclusions, and urgent abnormalities requiring immediate attention. As a result, models may achieve seemingly competitive scores while omitting content that is pivotal to patient safety. To address this limitation, several studies have proposed evaluation frameworks based on structured medical semantics. For example, some compute keyword retrieval rates over a limited set of lesion labels in chest radiography, whereas others quantify report content along dimensions such as severity, anatomical location, and imaging attributes in general radiology settings^30,31^. Nevertheless, these approaches are primarily designed for thoracic imaging or routine neuroimaging cohorts and struggle to represent the highly heterogeneous pathology spectrum of emergency brain CT, as well as emergency-specific findings tightly coupled to time-sensitive interventions. Furthermore, heterogeneity in data and clinical knowledge within emergency settings exacerbates the gap between model development and evaluation. Variations across institutions in scanning protocols, window settings, and reporting styles are difficult to fully capture using simple image-text pairing and conventional evaluation metrics^32,33^. Together, these limitations suggest that enabling multimodal large models to operate effectively in real-world emergency workflows requires a scenario-oriented redesign across dataset construction, model development, and evaluation design^34^.

In this study, we develop ERBrain, a multicenter multimodal framework for emergency brain CT interpretation designed to address key challenges in cross-institutional data heterogeneity, complex three-dimensional anatomy, and time-critical clinical decision-making. The framework begins with a standardized multicenter data pipeline that harmonizes volumetric CT examinations and corresponding radiology reports, enabling robust learning from whole-brain imaging inputs and structured Chinese clinical text. Building on this foundation, ERBrain integrates a pretrained three-dimensional visual backbone with a Chinese large language model within a unified autoregressive framework, facilitating joint learning of volumetric image understanding and report generation. To better capture clinically decisive abnormalities, we further introduce emergency-oriented optimization strategies that enhance sensitivity to critical signs and mitigate hallucinations or generation biases weakly supported by imaging evidence. ERBrain is designed to produce two clinically complementary outputs: emergency severity triage for rapid case prioritization and preliminary free-text report generation for radiologist review. To evaluate these capabilities, we propose FIES, a structured assessment framework spanning Feature, Impression, and Emergency Signs, which extends conventional text-based metrics by emphasizing semantic fidelity to emergency decision-making.

## Results

### Emergency brain CT clinical workflow and task formulation

Non-contrast emergency brain CT is the first-line imaging examination for patients with trauma and acute neurological emergencies after hospital admission. In routine practice (Fig. 1), patients are first assessed by emergency physicians, who determine the need for urgent imaging and subsequent management. Following CT acquisition, examinations enter the radiology workflow for formal interpretation and reporting. In high-volume settings, emergency physicians may directly review images for immediate decision-making. However, interpretive expertise and available attention can vary substantially, especially during busy shifts. Meanwhile, radiologists often handle multiple emergency examinations under significant time pressure, such that high-risk cases may not always be prioritized as early as clinically desirable when processed in chronological order.

**Fig. 1.**
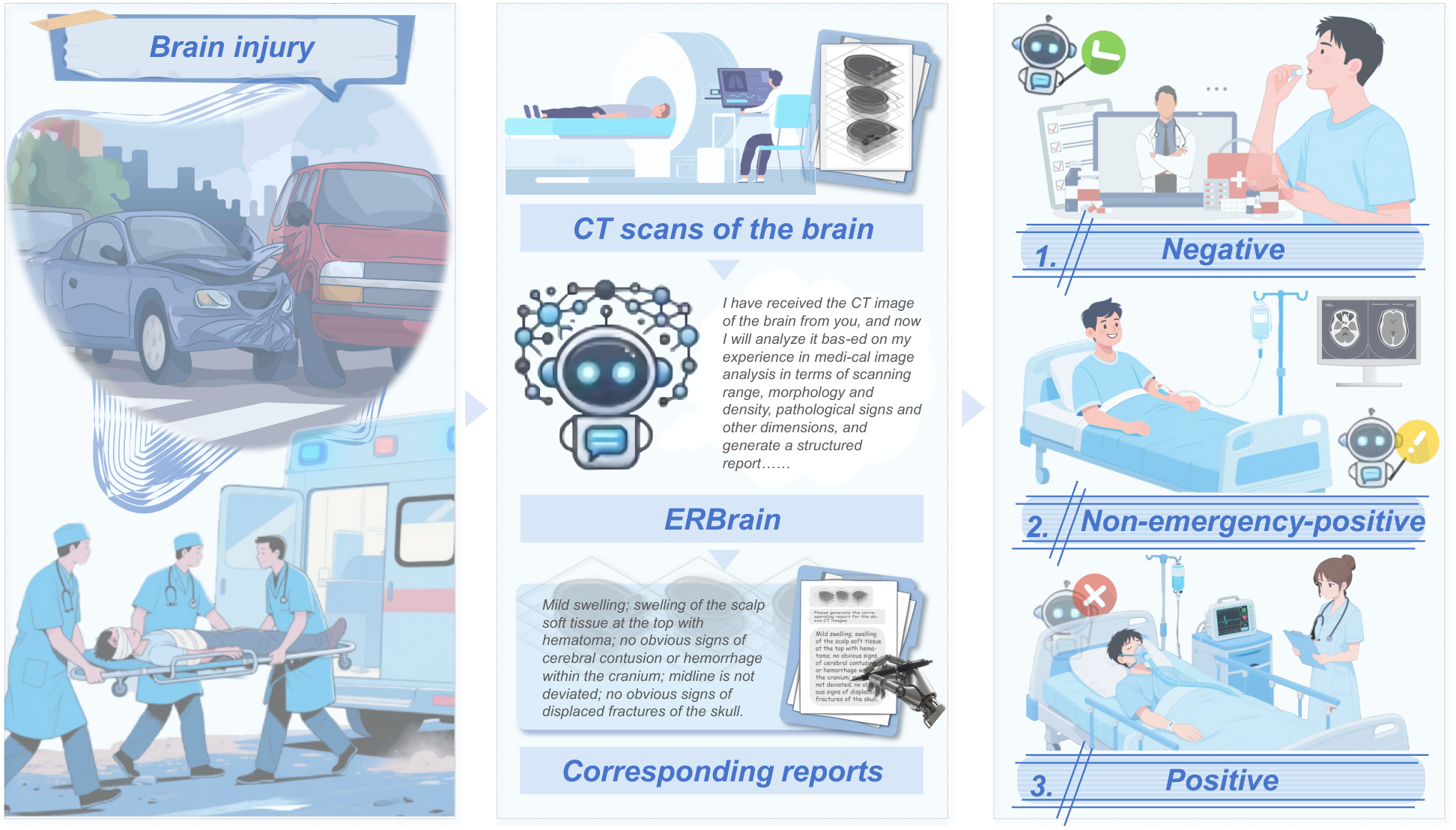
| Emergency brain CT clinical workflow and task formulation. a, Patients presenting with acute neurological symptoms or trauma are initially assessed in the emergency department and undergo non-contrast brain CT when clinically indicated. b, After image acquisition, the 3D brain CT volume is routed into the radiology workflow, where ERBrain analyzes the scan and generates a preliminary radiology report draft in Chinese. c, In parallel, ERBrain predicts a three-level emergency severity triage label as Negative, Non-emergency-positive, or Positive to support radiology worklist prioritization; Positive cases can then be communicated earlier to emergency physicians and relevant specialty teams while radiologists complete the formal report.

In this context, our system is designed to operate within the radiology workflow rather than as a stand-alone emergency department reader. Immediately after CT acquisition, the model analyzes the scan, assigns one of three emergency severity triage categories. These categories are Negative (no emergency abnormality), Non-emergency-positive (abnormalities present but not requiring immediate intervention), or Positive (critical emergency requiring urgent management). The model then generates a preliminary report draft. For cases flagged as urgent, the triage output and AI-generated draft are promptly relayed to emergency physicians to support early escalation and specialty consultation, while radiologists prioritize these cases for expedited review and completion of the formal report.

Building on this cognitive framework, we construct a complete task suite that spans emergency image interpretation from objective imaging understanding to clinical decision support. The suite is organized around three interrelated yet functionally independent core tasks: (1) extraction of structured semantics such as lesion characteristics, diagnostic impressions, and emergency signs; (2) automatic generation of emergency brain CT reports; (3) prediction of emergency severity levels. Collectively, these tasks delineate the essential capabilities required for an intelligent emergency radiology system to progress from pixel-level perception to decision-level reasoning. To systematically evaluate model performance in real emergency environments, we design a multidimensional experimental framework on multicenter data and validate it from four perspectives: report generation quality, completeness of semantic coverage, accuracy of critical condition recognition, and robustness under cross-hospital distribution shifts. The corresponding results are presented sequentially in the following sections.

### Structured semantic evaluation of emergency CT reports under FIES framework

To quantitatively assess the model’s expressive capacity at the level of clinical semantics, we conduct a systematic analysis of structured medical semantics in generated reports under the FIES framework. The three clinical semantic lexicons comprise 24 Feature keywords, 27 Impression keywords, and 71 Emergency Signs keywords (Fig. 2a). The keyword lexicon and structured evaluation dictionary used in the FIES framework are constructed exclusively from the training dataset, with no information from the validation or test sets involved in their development. Once defined, the lexicon is fixed prior to model evaluation and applied uniformly across all datasets to prevent potential information leakage. From the overall distribution, our model exhibits higher and more stable keyword hit frequencies across all three semantic dimensions, manifested as denser high-frequency hotspot regions. In contrast, M3D^19^, VoCo^35^, BrainGPT^15^, and RadFM^13^ generally show low frequency and dispersed distributions in the Feature and Emergency Signs dimensions, with only localized areas reaching moderate hit rates. These results indicate that our model provides a pronounced advantage in recognizing and expressing structured medical semantics, particularly emergency critical signs.

**Fig. 2.**
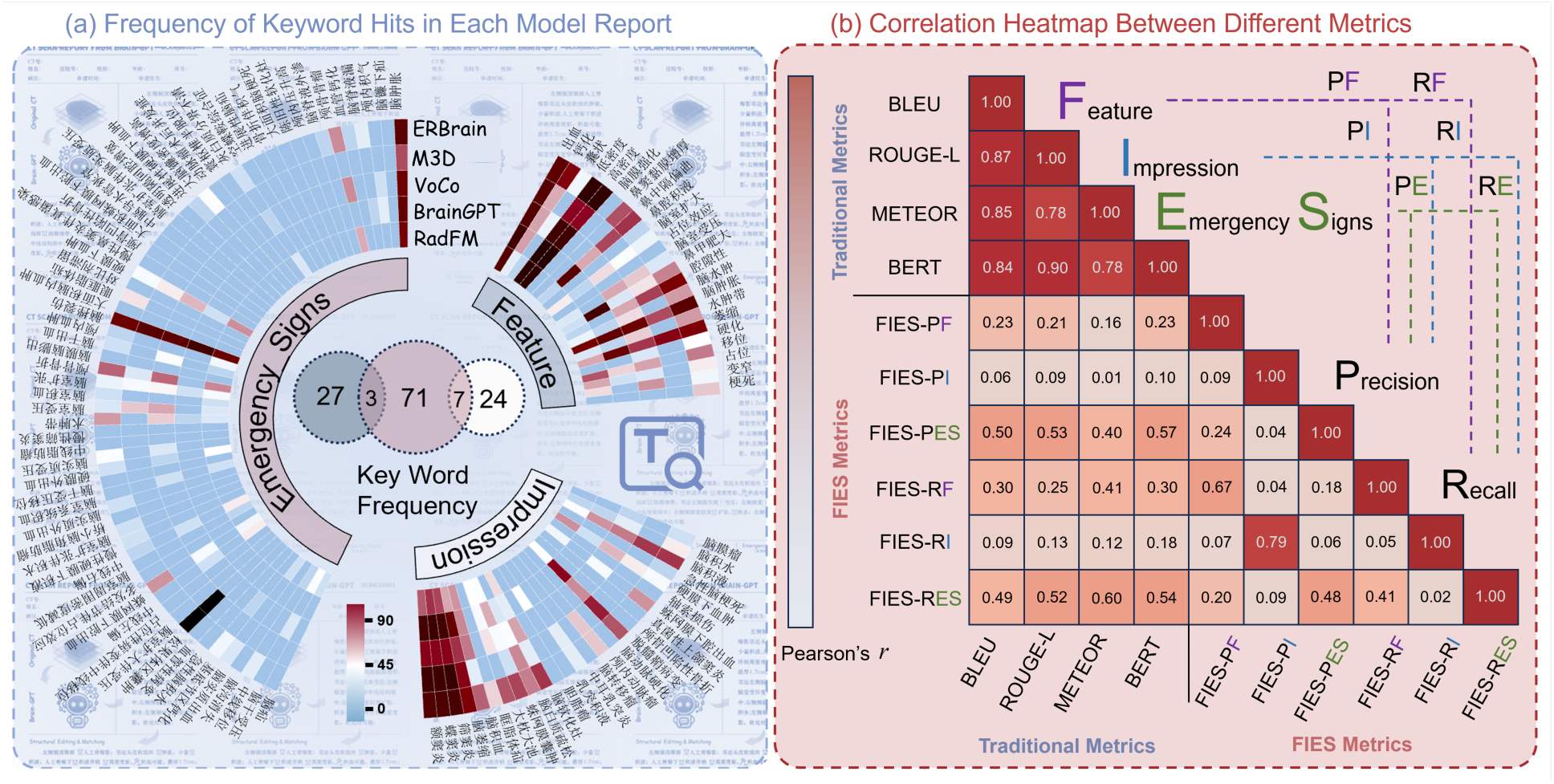
| Structured semantic evaluation of emergency brain CT reports under the FIES framework. a, Frequency of keyword hits in generated reports across the three FIES semantic dimensions, including Feature, Impression, and Emergency Signs. The circular heatmap summarizes per-keyword hit frequencies for ERBrain and baseline methods, with darker colors indicating higher hit frequency, and the center diagram shows the keyword set composition across the three lexicons (24 Feature keywords, 27 Impression keywords, and 71 Emergency Signs keywords). b, Pearson correlation heatmap between conventional report-generation metrics (BLEU, ROUGE-L, METEOR, and BERTScore) and FIES semantic metrics computed as precision and recall for Feature, Impression, and Emergency Signs, illustrating that conventional text similarity metrics correlate strongly with each other while showing weak correlations with decision-relevant FIES measures, thereby providing an orthogonal and clinically meaningful evaluation dimension for emergency radiology reporting.

To further interrogate the intrinsic relationships among evaluation metrics, we construct a Pearson correlation matrix between conventional language metrics and FIES semantic metrics (Fig. 2b). The results show strong correlations among conventional text similarity metrics. For example, ROUGE-L and BLEU achieve a correlation coefficient of approximately r = 0.78, and METEOR and BERTScore reach a comparable level, indicating that these metrics primarily capture linguistic form and surface matching. By contrast, correlations among the three FIES semantic dimensions are substantially lower, and the correlation between FIES-Impression and FIES-Emergency Signs is only r ≈ 0.04. This pattern reflects the independence of lesion description, diagnostic impression, and emergency signs in radiology writing.

More critically, there is almost no meaningful correlation between conventional language metrics and FIES metrics. For instance, the correlation between BLEU and FIES-Impression is only r ≈ 0.06, and the correlation between BERTScore and FIES-Impression is about r ≈ 0.01. This phenomenon indicates that even when the generated text is highly similar to the reference report at the linguistic level, it does not necessarily cover the medical semantics that are essential for emergency decision-making. This gap explains why some baseline models can appear acceptable under conventional metrics while still exhibiting clinically salient omissions.

### Structured semantic consistency and cross-metric performance analysis

After establishing that the model provides stronger semantic coverage, we further conduct a systematic comparison of generation quality across models under the FIES dimensions from two perspectives, namely metric distributions and semantic consistency. The case-level metric distribution analysis (Fig. 3a) summarizes overall differences in generation quality across four conventional language metrics and three FIES semantic metrics. Our model achieves higher medians and more concentrated distributions on all conventional metrics, reaching statistically significant differences relative to all baseline methods. Under the FIES semantic metrics, the model likewise attains the highest scores across Feature, Impression, and Emergency Signs, with particularly pronounced gains for Emergency Signs. This gain is reflected by an upward-shifted distribution and a marked reduction in outliers, indicating advantages in both recall and stability for emergency critical signs. Across the seven evaluation metrics, our model ranks first in nearly all cases, with only a slightly lower BERTScore than M3D. This gap likely arises because BERTScore emphasizes surface-level lexical similarity, whereas our model prioritizes emergency-critical and clinically salient semantics through structured, risk-focused generation.

**Fig. 3.**
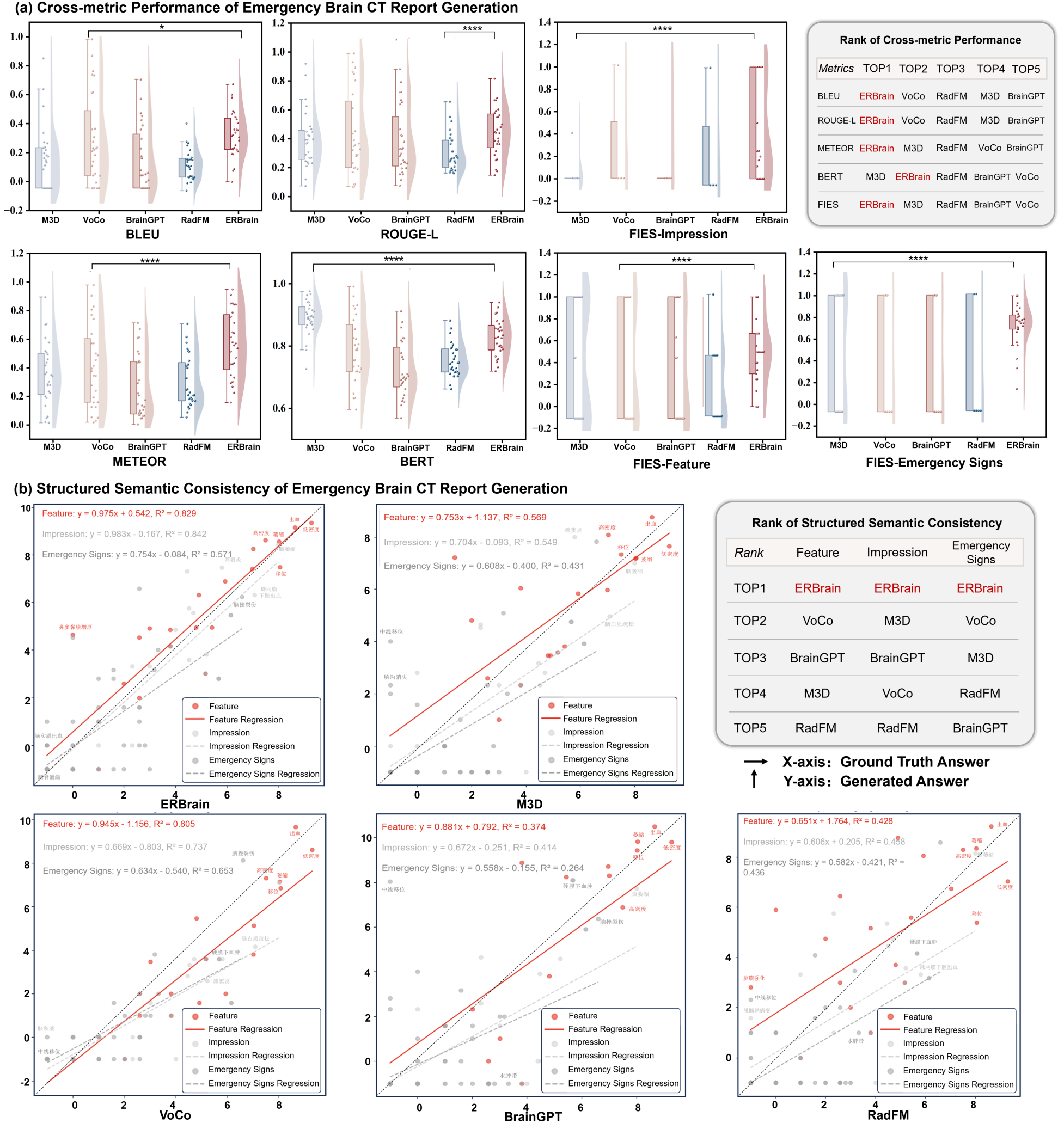
| Cross-metric performance and structured semantic consistency of emergency brain CT report generation. a, Case-level distributions of conventional report generation metrics and FIES semantic metrics for emergency CT report generation across ERBrain and baseline methods. Each panel shows the sample-wise metric distribution, and the ranking table summarizes the overall top-performing methods under each metric. b, Consistency analysis between generated reports and reference reports in the structured semantic space. For each method, scatter plots relate the keyword counts extracted from generated reports to the corresponding counts from ground-truth reports for Feature, Impression, and Emergency Signs, with fitted regression lines quantifying semantic proportionality. The ranking table summarizes consistency performance across the three semantic categories.

To evaluate the model’s stability in structured semantic space, we further assess the quantitative consistency between generated reports and reference reports across the three FIES semantic categories (Fig. 3b). By fitting a log regression over keyword counts, our model exhibits high consistency across Feature, Impression, and Emergency Signs. The regression slopes for Feature and Impression are 0.975 and 0.983, respectively, with corresponding R^2^ values exceeding 0.82; even in the most challenging Emergency Signs dimension, the model maintains a regression slope of 0.754 and an R^2^ of 0.571. In contrast, most baseline methods markedly underestimate emergency signs, with generally low slopes and R^2^ values, indicating that their expression of critical information is neither sufficient nor stable. These results underscore that consistency in structured semantic space is closely associated with clinical usability. Beyond overall metric leadership, our model more closely approximates real radiology writing behavior in semantic proportionality and expression stability, thereby exhibiting greater application potential for emergency brain CT report generation.

### Case-level and in-distribution evaluation of emergency brain CT report generation

Using multiple representative emergency brain CT cases (Fig. 4), we perform sentence-by-sentence comparative analyses between model-generated reports and reference reports. The case-level results show that our model maintains high stability in content organization, extraction of key imaging findings, and construction of diagnostic reasoning. For example, in acute intracranial hemorrhage cases, some baseline models provide vague or nonspecific descriptions and fail to specify hemorrhage location, extent, and potential risks. By explicitly localizing abnormal hyperdense foci and jointly modeling their distribution and morphologic features, our model produces structured descriptions that support a clearer diagnostic impression consistent with emergency radiologic reasoning. Keyword-hit visualization further illustrates reference-based FIES term coverage. Across the three FIES dimensions, ERBrain matched 7 of the 9 FIES terms identified in the reference report, compared with 1/9 for M3D, 2/9 for VoCo, and 5/9 each for BrainGPT and RadFM. These keywords cover critical imaging signs such as “高密度灶” (“hyperdense foci”) and “脑实质密度异常” (“abnormal parenchymal density”), reflecting the model’s ability to consistently recognize and express core lesion information. In the Impression dimension, the model generates explicit diagnostic conclusions, accurately identifying conditions such as “急性颅内出血” (“acute intracranial hemorrhage”). In the Emergency Signs dimension, it captures high-risk findings such as “中线移位” (“midline shift”) and clearly articulates them in the generated text.

**Fig. 4.**
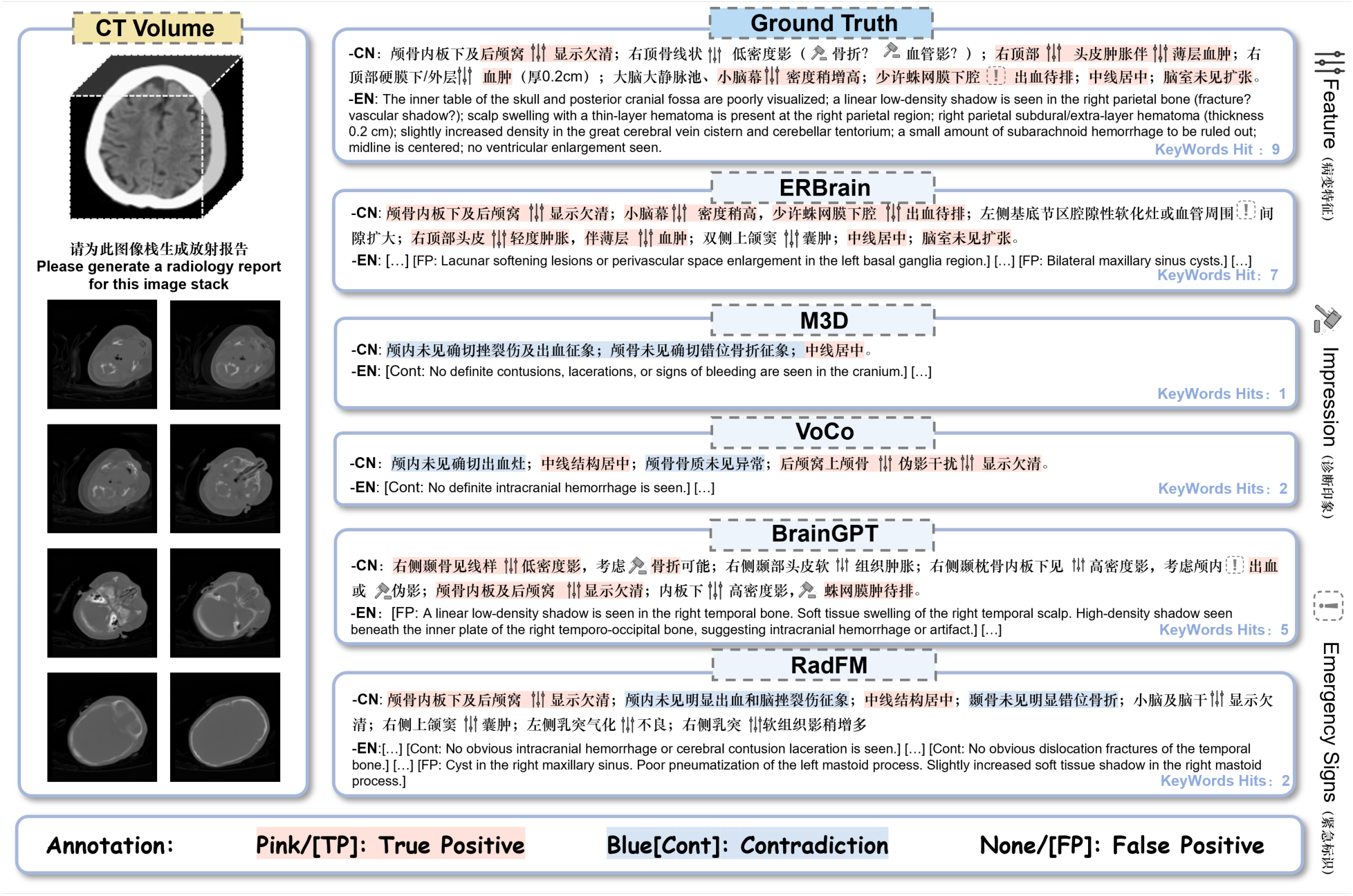
| Case-level comparison of emergency brain CT report generation, FIES keyword coverage, and error analysis. A representative emergency brain CT case is shown with the input CT volume and the instruction prompt on the left, and the reference report and generated reports on the right. From top to bottom, outputs are presented for the reference report, ERBrain, and different baseline methods. For each report, highlighted terms indicate matches FIES keywords, and the keyword-hit count is shown for the case, enabling direct comparison of clinical semantic coverage across Feature, Impression, and Emergency Signs. Color annotations indicate reference-report-based phrase-level comparisons, where pink denotes findings consistent with the reference report, blue denotes statements that contradict the reference report, and unhighlighted text denotes generated findings absent from the reference report.

The case-level observations are consistent with large-scale distribution quantitative evaluation trends (Fig. 5). On in-distribution data from West China Hospital and Mianyang Central Hospital, our model achieves a BLEU of 0.2603 and a METEOR of 0.4458, clear improvements over the strongest baseline. It also ranks second on ROUGE-L and BERTScore. More importantly, under FIES metrics that reflect clinical semantic coverage, the model significantly outperforms all baseline methods, with FIES-Avg reaching 0.5465, representing a 20.8% improvement over the strongest baseline, thereby demonstrating a substantive advantage in capturing emergency critical semantics.

**Fig. 5.**
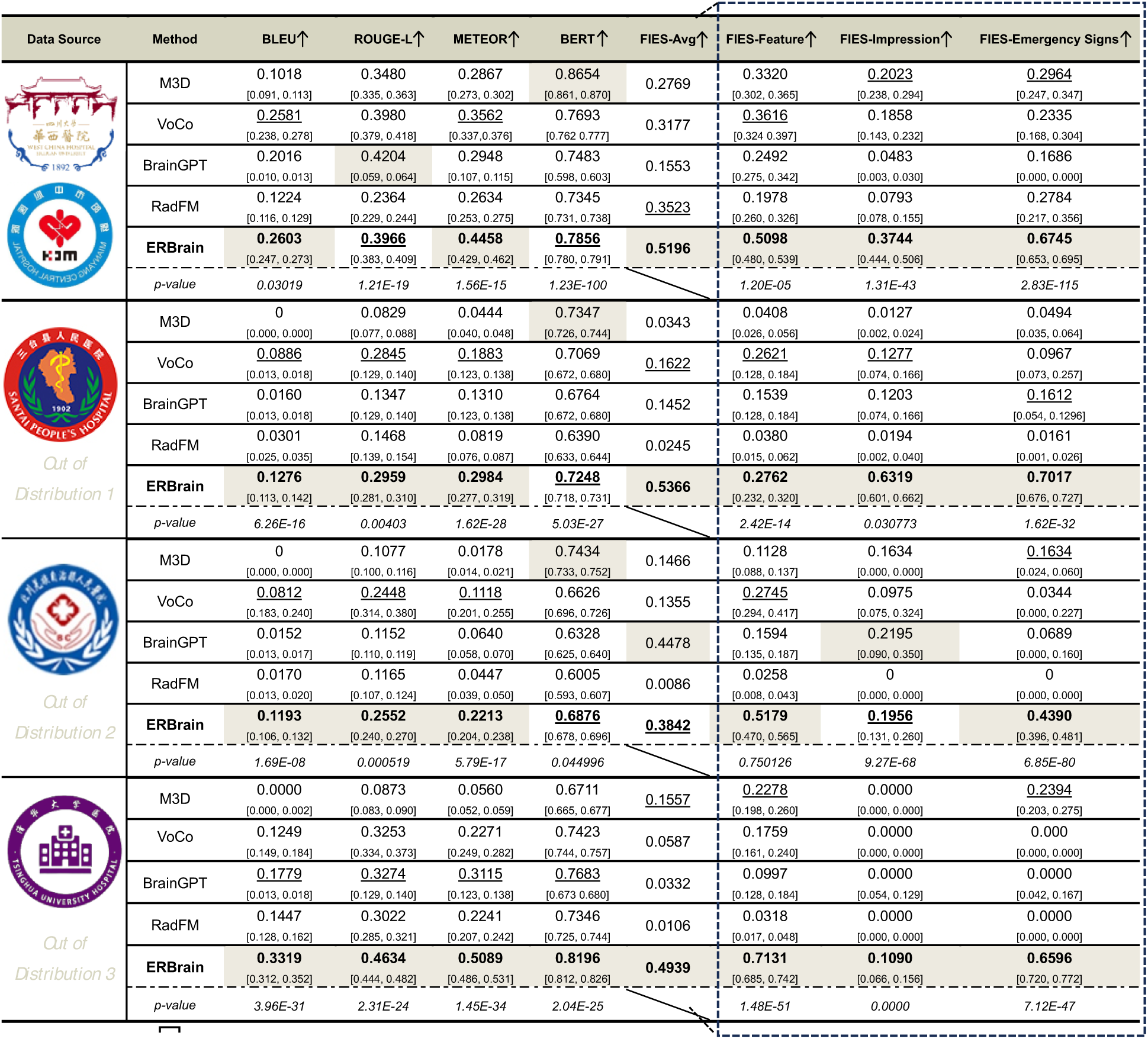
| Quantitative comparison of emergency brain CT report generation across hospitals. ERBrain is compared with representative multimodal report generation baselines on the in-distribution test set and three out-of-distribution hospital cohorts. Report quality is evaluated using conventional text similarity metrics (BLEU, ROUGE-L, METEOR, and BERTScore) together with the proposed FIES metrics (FIES-Avg, FIES-Feature, FIES-Impression, and FIES-Emergency Signs), with higher values indicating better performance. Each cell reports the point estimate, and the bracketed range reports the corresponding 95% CI of the score. The p-value row summarizes the statistical significance of ERBrain relative to the strongest competing baseline under the same cohort setting.

### Emergency severity triage and diagnostic consistency analysis

In addition to report generation, we further evaluate the model’s performance on emergency severity triage, in comparison with baseline methods RIDL^36^, TransLiver^37^, SC-Net^38^, and LMTTM^39^. On in-distribution data (Fig. 6a), our model achieves the highest values across Accuracy (0.9434), Balanced Accuracy (0.9403), Macro-Precision (0.9401), Macro-Recall (0.9403), Macro-F1 (0.9402), Macro-Specificity (0.9667), and Macro G-mean (0.9491). Compared with the strongest baseline method RIDL, the model achieves an absolute improvement exceeding 0.39 in Balanced Accuracy (0.9403 vs. 0.5453), an absolute improvement exceeding 0.46 in Macro-Recall (0.9403 vs. 0.4780), and an improvement exceeding 0.40 in Macro G-mean (0.9491 vs. 0.5464), reflecting more balanced recognition across the three triage categories. These aggregate metrics are macro-averaged across classes and should not be interpreted as Positive-class sensitivity; the Positive-class recall was 90.1% (192 of 213 cases) in the in-distribution cohort.

**Fig. 6.**
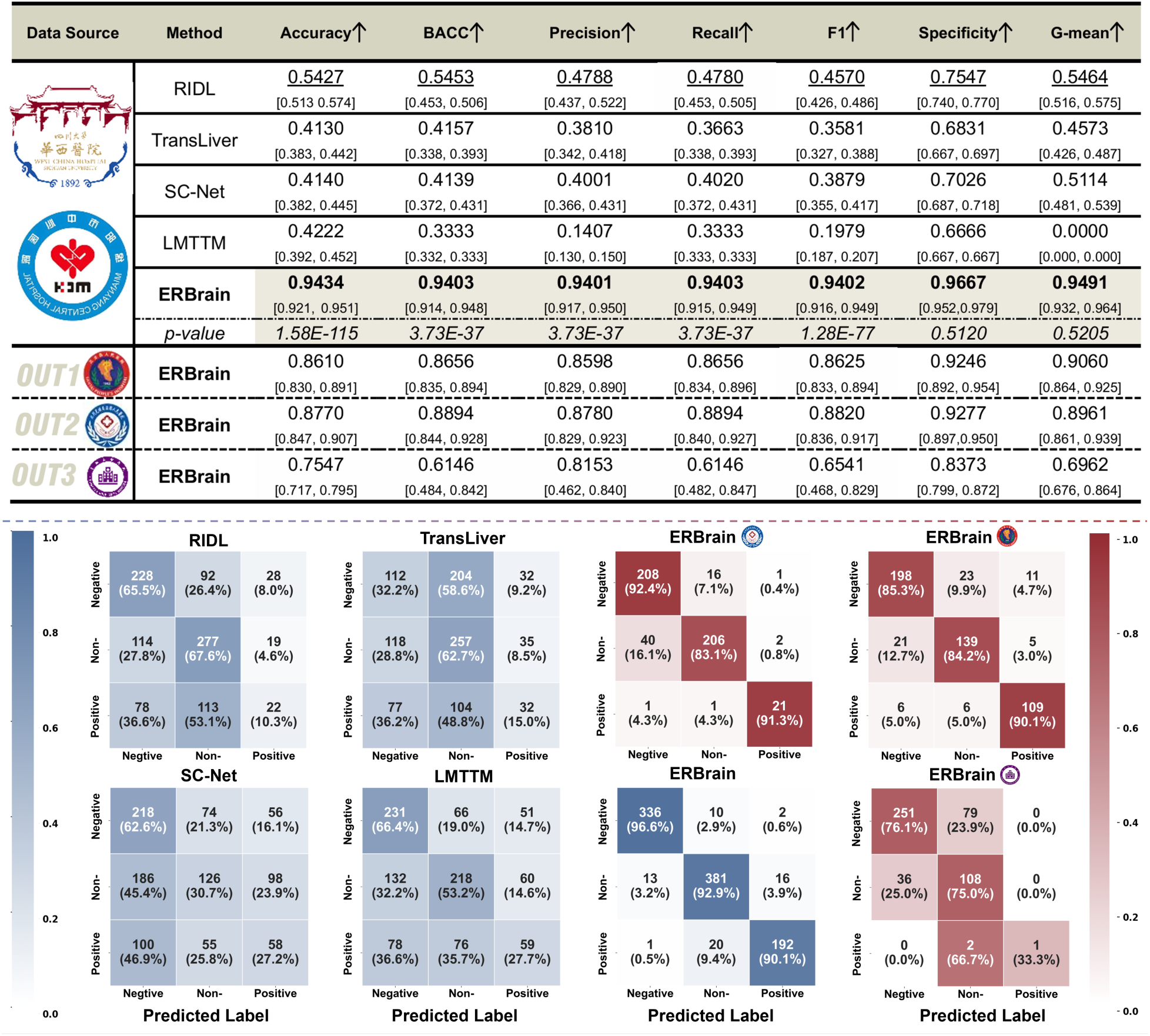
| Three-level emergency severity triage performance and confusion matrices. Top, ERBrain is compared with triage baselines on the in-distribution test set, and further evaluated on three out-of-distribution hospital cohorts. Performance is reported with Accuracy, BACC, Precision, Recall, F1, Specificity, and G-mean. Each cell reports the point estimate, and the bracketed range reports the corresponding 95% CI of the score. The p-value row summarizes the statistical significance of ERBrain relative to the strongest baseline on the in-distribution test set. Bottom, normalized confusion matrices visualize label-wise performance for the compared methods and for ERBrain under distribution shift, with rows as ground-truth classes and columns as predicted classes.

Meanwhile, confusion matrix analysis (Fig. 6b) further characterizes class-level error distributions. The normalized confusion matrix shows that our model achieves strong and well-balanced classification performance across the three categories. The diagonal proportions for Negative, Non-emergency-positive, and Positive are 96.6%, 92.9%, and 90.1%, respectively, indicating relatively consistent and robust discrimination across emergency severity triage classes. Inter-class misclassification rates were low overall; however, 21 of 213 Positive cases were classified as either Non-emergency-positive or Negative, underscoring the need for lesion-level characterization of these false-negative cases. In contrast, most baseline models exhibit pronounced error diffusion across classes. In particular, TransLiver and SC-Net produce a substantial number of false-positive assignments in normal cases, leading to markedly reduced overall specificity.

### Generalization under cross-hospital distribution shifts

In real emergency applications, substantial differences across medical institutions are common in scanning protocols, scanner models, image quality, and case composition; therefore, performance stability under cross-hospital distribution shifts is a key indicator of clinical deployability. We conduct a systematic evaluation of report generation and emergency severity triage in two external out-of-distribution validation cohorts from Santai County People’s Hospital and Beichuan Qiang Autonomous County People’s Hospital. We also include a low-prevalence external stress-test cohort from Tsinghua University Hospital. In emergency brain CT report generation (Fig. 5), baseline models generally exhibit pronounced performance degradation in out-of-distribution settings. For example, at Santai County Hospital, BLEU for VoCo and M3D drops to 0.0886 and 0, respectively. Consistent degradations are also observed in METEOR and BERTScore across multiple baselines, indicating that language generation performance is difficult to sustain under cross-institutional distribution shifts. By contrast, our model achieves comparatively favourable report-generation performance across the three external cohorts. Its BLEU reaches 0.1276, 0.1193, and 0.3319; ROUGE-L reaches 0.2959, 0.2552, and 0.4634; and METEOR reaches 0.2984, 0.2213, and 0.5089. These represent the best or tied-best values under each hospital condition, indicating greater relative robustness than the evaluated baseline methods under cross-institutional distribution shifts.

More importantly, under structured semantic metrics that better reflect clinical usability, our model maintains stable coverage of core medical semantics despite distribution shifts. Specifically, FIES-Feature reaches 0.2237, 0.5160, and 0.7131 across the three hospitals, and FIES-Emergency Signs, which is particularly critical for emergency decision making, remains at 0.5721, 0.2848, and 0.6596, without systematic omission of critical signs. In contrast, most baseline models show substantial declines in this dimension, and some approaches degrade to near failure, indicating that they cannot reliably support emergency risk assessment in cross-institutional environments. To assess semantic preservation during DeepSeek-V3 report normalization, we compared prespecified clinical keyword sets between the original and normalized reports in the Santai cohort (n = 518; Supplementary Table 1). Micro keyword retention recall was 85.1%, micro preservation precision was 96.2%, and exact keyword-set agreement was observed in 422 of 518 report pairs (81.5%), indicating that normalization generally preserved the measured clinical keywords but was not semantically lossless.

In the emergency severity triage task, out-of-distribution settings further highlight robust differences across models (Fig. 6). Because conventional methods deteriorate substantially on external data, they are not included in the quantitative comparison. ERBrain maintains favourable classification performance in the Santai and Beichuan external validation cohorts, achieving accuracies of 0.8610 and 0.8770 and G-mean scores of 0.9059 and 0.8961, respectively. In contrast, performance is lower in the Tsinghua low-prevalence stress-test cohort, with an accuracy of 0.7547 and a G-mean of 0.6962. These results indicate a favourable balance between sensitivity and specificity in the two external validation cohorts. Confusion matrix analysis shows Positive-class recall values of 90.1% in Santai and 91.3% in Beichuan. In the Tsinghua stress-test cohort, only one of three Positive cases was correctly classified; therefore, the corresponding class-specific estimate is considered exploratory and is not interpreted as stable evidence of Positive-class external generalization.

### Statistical Analysis

All statistical analyses are performed at the patient level using paired comparisons between models evaluated on the same cases. The p-value rows summarize the statistical significance of differences between ERBrain and the strongest non-ERBrain baseline for each cohort and metric. For report-generation metrics with case-level paired scores, including BLEU, ROUGE-L, METEOR, BERTScore, and FIES-based metrics, significance is assessed using two-sided Wilcoxon signed-rank tests. ERBrain shows significant differences from the strongest competing baseline across cohorts for the conventional report-generation metrics. For example, in the in-distribution cohort, P values are 3.02×10^-2^ for BLEU, 1.21×10-^19^ for ROUGE-L, 1.56×10^-15^ for METEOR, and 1.23×10^-100^ for BERTScore. Most FIES-based comparisons are also significant, supporting that the observed improvements in clinically relevant semantic coverage are unlikely to be attributable to random variation alone, although significance varied across semantic dimensions in the external cohorts (Fig. 5).

For cohort-level triage metrics, 95% confidence intervals (CIs) and P values are estimated using paired nonparametric bootstrap resampling at the patient level with replacement over 2,000 iterations. ERBrain significantly outperforms the strongest competing baseline on the prespecified primary triage endpoint of overall accuracy (P = 1.58×10^-115^), as well as on balanced accuracy and macro-F1 (P = 3.73×10^-37^ and 1.28×10^-77^, respectively), whereas differences in macro-specificity and macro G-mean are not statistically significant (P = 0.5120 and 0.5205, respectively). These findings indicate that the superior retrospective triage performance of ERBrain is supported not only by higher point estimates, but also by formal statistical significance in the primary and key secondary accuracy-related metrics. The prespecified primary endpoints are FIES-Avg for report generation and overall accuracy for emergency severity triage (Fig. 6).

### Calibration Analyses

Calibration analysis demonstrates that our model maintained well-calibrated probability estimates in the internal cohort (Fig. 7). The calibration curves for the Negative, Non-emergency-positive, and Positive classes all closely followed the identity line, with low Brier scores of 0.023, 0.048, and 0.031, respectively (n = 348, 410, and 213), indicating good agreement between predicted probabilities and observed frequencies. Across external hospitals, although some center-level variation is observed, the model largely maintains calibration consistency. For the Santai County People’s Hospital cohort, Brier scores are 0.097, 0.089, and 0.058 for the three classes (n = 232, 165, and 121), respectively. In the Qiang Autonomous County People’s Hospital cohort, Brier scores are 0.079, 0.096, and 0.027 (n = 225, 248, and 23), respectively. The Tsinghua University Hospital cohort exhibited greater calibration deviation, with Brier scores of 0.204, 0.151, and 0.030 for the three classes (n = 330, 144, and 3). This deviation is likely due to a highly imbalanced case mix and the extremely limited number of positive cases.

**Fig. 7.**
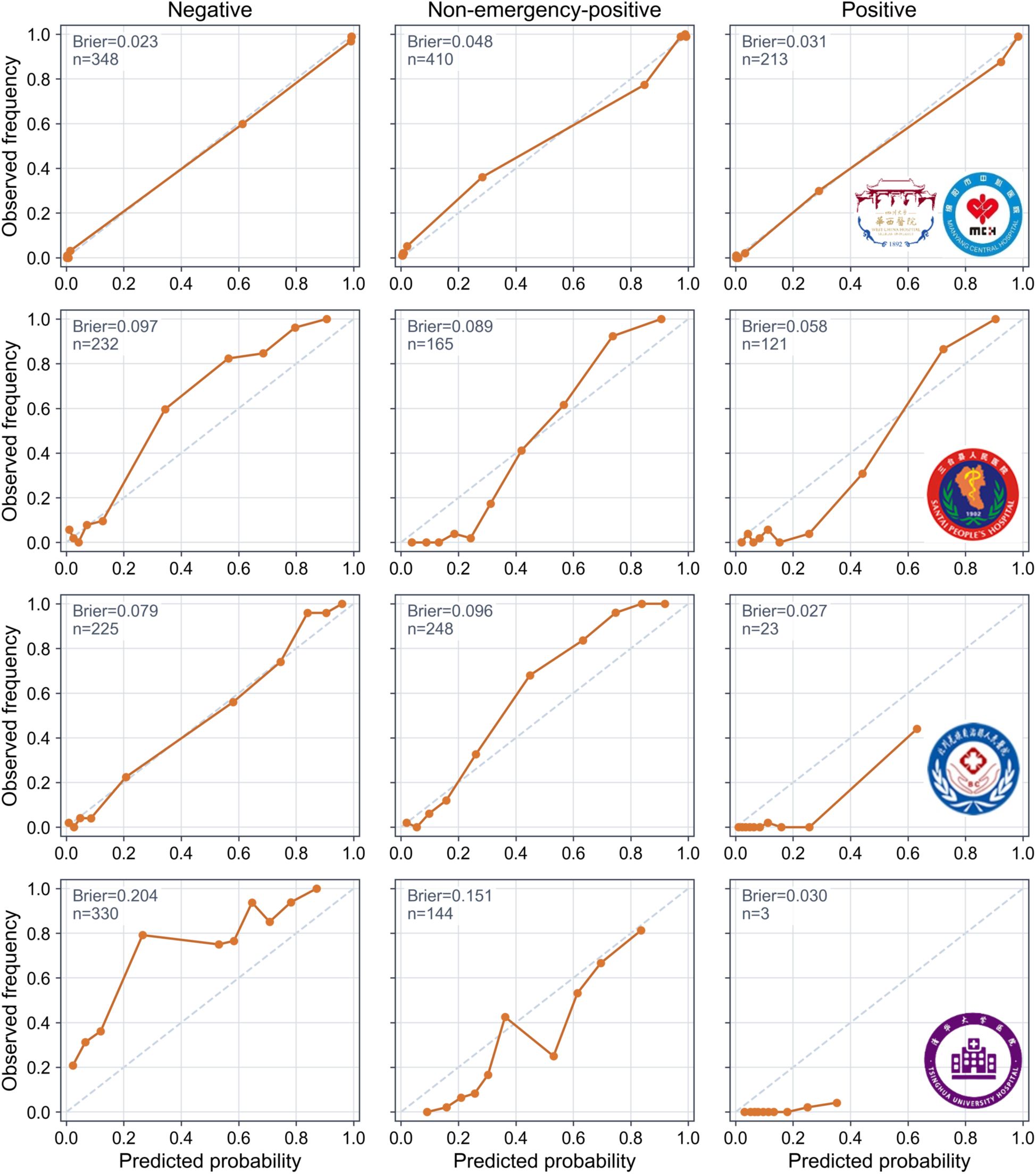
| Calibration of the primary classification task across the in-distribution and out-of-distribution cohorts. Calibration plots are shown for the three classification categories, negative, non-emergency-positive and positive, across the in-distribution cohort and three out-of-distribution cohorts. The x axis indicates predicted probability and the y axis indicates observed frequency. The dashed diagonal denotes perfect calibration. Orange lines show the empirical relationship between predicted probabilities and observed event frequencies within probability bins. The Brier score and the number of cases are shown in each panel.

## Discussion

This study presents a multimodal large model system tailored to emergency brain CT, which, within a unified framework, enables automated radiology report generation from three-dimensional brain CT volumes while simultaneously determining the emergency severity tier of intracranial disease. Systematic evaluation on multicenter retrospective clinical data shows that the proposed system maintains consistent advantages on conventional text generation metrics and substantially improves the recall and completeness of high-risk emergency signs under our proposed FIES emergency semantic metrics. On the emergency triage task, it further achieves higher sensitivity and stronger risk discriminability. These findings indicate that a multimodal large model can be explicitly anchored to the high-risk, strongly time-critical setting of emergency brain CT, with both image-to-text quality and triage decision consistency reinforced within the training objectives and evaluation protocol. Under this design, its capability evolves beyond merely restating imaging observations toward a more clinically actionable level that supports emergency triage and clinical decision making.

Beyond model-level performance, ERBrain has important implications for emergency workflow optimization, particularly in the context of emerging CT-first paradigms. In conventional Advanced Trauma Life Support workflows, patients are typically routed through the trauma bay prior to imaging, introducing non-negligible delays during the critical early phase of care. Although CT-first strategies have been increasingly advocated to accelerate diagnosis, their practical adoption has been constrained by the lack of immediate expert interpretation. In this regard, ERBrain directly addresses the interpretation latency bottleneck by aligning diagnostic speed with image acquisition. Specifically, on a single NVIDIA A800 GPU with batch size 1, the end-to-end inference latency is 0.1802 s per case, including approximately 0.02 s for preprocessing, which is negligible compared with patient transport and workflow transition times. This near real-time capability enables preliminary triage decisions to be generated at the point of imaging, thereby supporting more streamlined patient routing and potentially reducing unnecessary transfers. Consequently, ERBrain provides a feasible technological foundation for integrating rapid, automated triage into CT-first emergency pathways, while remaining compatible with clinician-in-the-loop decision making.

Unlike prior studies that focus solely on image classification or treat radiology report generation as a standalone task, our approach jointly models report generation from three-dimensional brain CT and emergency severity stratification on a shared multimodal backbone. This design more faithfully mirrors the real workflow of radiologists in emergency settings. Experimental results demonstrate that the triage branch serves not only as an independent source of discriminative supervision but also as a structural constraint during generation. This constraint reduces vague and overly conservative template-based descriptions and instead encourages outputs that contain explicit diagnostic impressions and emergency alerts that are coherent with the predicted triage category. From a clinical perspective, this joint modeling paradigm ensures that generated reports are not isolated textual artifacts, but inherently carry risk signals aligned with emergency triage logic. This alignment enables direct use for priority ranking and triage decision support within emergency imaging workstations^40,41^. These results suggest that, in high-risk medical scenarios, synergistically coupling image to text generation with an explicit downstream stratification objective is a key pathway toward improving the clinical usability and decision value of multimodal large models.

Another key contribution of this study is the introduction of FIES metrics for emergency brain CT reporting, designed to address blind spots of general natural language generation metrics, including BLEU, ROUGE, METEOR, and BERTScore, within clinical contexts. We decompose reports into structured keywords across three categories: lesion characteristics, diagnostic impressions, and emergency signs. Under this decomposition, certain model variants that show only marginal gains or even parity on conventional metrics achieve substantially higher precision and recall on the emergency signs dimension of FIES, which directly reflects the coverage of high-risk information that clinicians most prioritize. Conversely, some models attain relatively high scores on conventional metrics yet are revealed by FIES to omit or misstate critical signs, suggesting that reliance on general NLP metrics alone may overestimate clinical reliability. This observation aligns with recent discussions on structured keyword-based evaluation and LLM-as-a-judge paradigms. Such work underscores that radiology report assessment should emphasize interpretable dimensions directly linked to diagnostic and treatment decisions, rather than focusing solely on n-gram overlap or generic semantic similarity^42,43^. In this sense, FIES not only provides a more clinically aligned measurement tool for our study but also offers a transferable design blueprint for building organ-specific and scenario-specific evaluation benchmarks.

In designing the training objectives, we integrate a critical information loss, a dynamic retrospective learning loss, and the base language modeling loss to form a joint optimization strategy tailored to emergency settings. The critical information loss dynamically upweights high-risk tokens, such as midline shift, brain herniation, and ventricular compression. This upweighting concentrates gradient updates on semantics that are tightly coupled to patient outcomes and improves sensitivity to emergency signs even under limited annotation^44^. The dynamic retrospective learning loss constrains alignment between global visual features extracted from three-dimensional CT and features derived from ground truth report text. This constraint mitigates common issues in multimodal generation, including template-like outputs and hallucinated content, and reduces statements that conflict with imaging evidence in clearly positive cases^45^. Building upon these objectives, the knowledge-augmented prompting strategy allows the model to dynamically invoke guideline-level and review-level knowledge when encountering complex or atypical cases. This embeds imaging findings within a richer clinical context and reduces over-reliance on statistical co-occurrence patterns in the training corpus. Taken together, this integrated optimization strategy, combining emergency vocabulary strengthening, visual retrospective constraints, and external knowledge injection, provides a replicable mechanistic paradigm for multimodal system design in other high-risk specialty settings^46,47^.

Importantly, retrospective cross-hospital evaluation showed that the model maintained favourable performance in the Santai and Beichuan external validation cohorts, whereas performance declined in the Tsinghua low-prevalence stress-test cohort. Data engineering procedures, including image preprocessing, text structuring, and report normalization, may partially mitigate distribution shifts arising from differences in scanning protocols and reporting practices. These results suggest that, for organ and scenario-specific clinical tasks, data governance and model design should be treated as an inseparable whole, and a strong multimodal backbone alone is insufficient to guarantee clinical usability. In parallel, the domestically developed Chinese large language model adopted in this study achieves stable and interpretable performance in emergency neuroimaging after targeted visual alignment, emergency semantic strengthening, and retrieval augmented knowledge injection. This result highlights the practical potential and controllable deployment costs of domestic models in specialized medical tasks. Furthermore, our emergency brain CT keyword library and the FIES evaluation suite can be viewed as an open infrastructure. This infrastructure may be combined with other vision language models to serve as a unified training and evaluation substrate for brain imaging tasks. Moreover, the proposed framework is inherently language- agnostic and can be extended to international clinical settings by generating reports in the native language and subsequently applying standardized translation, enabling consistent emergency reasoning across diverse linguistic environments.

Despite these advances, several limitations should be acknowledged. First, the current labels for emergency severity are primarily derived from a three-tier scheme used at a single center and do not yet support fine-grained differentiation of risk according to specific therapeutic time windows, intervention intensity, or monitoring requirements. This limits the granularity with which model outputs can be aligned to real emergency management pathways^48^. Second, our validation focuses on report generation and emergency severity classification. We have not yet systematically assessed finer-grained tasks such as abnormality localization, lesion detection, or segmentation, which restricts direct applicability to scenarios including preoperative planning and longitudinal disease monitoring^49,50^. Third, the current system relies on non-contrast head CT alone. This CT-only design imposes an inherent ceiling on utility for emergency department decision-making. This ceiling arises because urgency in borderline or clinically complex cases is often determined not only by imaging findings, but also by neurologic examination findings, anticoagulation status, symptom onset timing, vital signs, laboratory results, and prior imaging. Finally, although retrospective and in-silico validation provides important preliminary evidence, performance observed under these conditions may overestimate effectiveness in real-world deployment. Clinical implementation may be affected by dataset shift, institutional workflow variation, differences in patient populations across hospitals, and human-factor challenges such as alert fatigue, inappropriate trust calibration, and ambiguity in medico-legal responsibility.

Future work will address these limitations along several directions. We will introduce more fine-grained and clinically grounded risk stratification labels that better reflect actionable emergency management needs, including differences in treatment urgency, monitoring intensity, and intervention pathways. We also plan to extend the current framework beyond report generation and severity classification by incorporating downstream tasks such as abnormality localization, lesion detection, and segmentation into a unified modeling pipeline. To overcome the ceiling imposed by CT-only inputs, an important next step will be to investigate multimodal integration, particularly the addition of brief structured clinical context to imaging inputs. We will then evaluate whether such multimodal information improves triage performance, calibration, and safety. Until such multimodal evidence is established, the intended use of the current system should be regarded as a radiology worklist prioritization and triage assistant rather than a comprehensive emergency department decision support system. In parallel, following the DECIDE-AI framework and major society recommendations, future evaluation should move beyond retrospective validation toward prospective multicenter studies and robust randomized controlled trials, with particular emphasis on patient-centered outcomes, workflow efficiency, and real-world clinical effectiveness. Continuous monitoring, model updating, and governance mechanisms will also be essential to support safe and reliable deployment over time.

## METHODS

### Multicenter emergency data and standardized preprocessing

In this study, we construct a 3D CT text dataset based on multicenter emergency brain CT images and reports to retrospectively evaluate model generalization under cross-hospital distribution shifts in multicenter clinical data (Supplementary Fig. 1). Reporting of this retrospective multicenter study is informed by TRIPOD-LLM-relevant items for healthcare studies involving large language models, particularly with respect to study design, inference reproducibility, prompt transparency, human oversight, and intended use description. The dataset comprises more than 10,000 cases, covering real emergency examinations from multiple medical institutions and spanning the full age spectrum, including pediatric, young to middle-aged, and older adult patients (Supplementary Fig. 2). In the experimental design, we partition the data into in-distribution (ID) and out-of-distribution (OOD) subsets according to the originating institution. Cases from West China Hospital (n = 5,648) and Mianyang Central Hospital (n = 4,370) were used as in-distribution data. Cases from Santai County People’s Hospital (n = 518) and Beichuan Qiang Autonomous County People’s Hospital (n = 496) served as external out-of-distribution cohorts for cross-institutional validation. Tsinghua University Hospital (n = 477) was analysed separately as a low-prevalence external stress-test cohort because it contained only three Positive cases. Santai and Beichuan are county-level institutions with scanner configurations, acquisition protocols, and case compositions that differ from those of the in-distribution centres, whereas the Tsinghua cohort primarily evaluates performance under a pronounced prevalence and class-composition shift. Accordingly, class-specific estimates for the Positive category in the Tsinghua cohort are treated as exploratory and are not interpreted as stable external-validation estimates. Scanner models and acquisition parameters varied across the five participating centers (Supplementary Table 2).

To reduce distributional discrepancies across centers in both imaging conditions and textual expression, and to provide consistent inputs for subsequent multimodal modeling, we apply a unified standardized preprocessing pipeline to images and reports, respectively (Fig. 8). Representative emergency brain CT examples are provided for each cohort and triage category (Supplementary Fig. 3). To address heterogeneity across centers in scanning protocols, slice thickness distributions, and image quality, all raw CT volumes are processed with z axis resampling, removal of non-tissue slices, spatial normalization, and resolution standardization to 32×256×256. We then apply HU windowing truncation followed by linear normalization. To accommodate slice thickness and noise differences in out-of-distribution data, we additionally perform center reconstruction and intensity correction. These steps are intended to reduce variability arising from differences in acquisition quality, slice thickness, noise, and scanner-specific intensity characteristics across centers. Notably, the present study does not include a dedicated failure-mode analysis for severely degraded inputs, such as marked motion artifacts, incomplete series, or extreme noise; such cases would still require radiologist review in the intended workflow. The resulting volumes are saved in float32 format as unified “.npy” files and used as the unified visual inputs to the model. On the text side, all original radiology reports are cleaned, normalized for medical terminology, segmented at the sentence level, and structurally parsed.

**Fig. 8.**
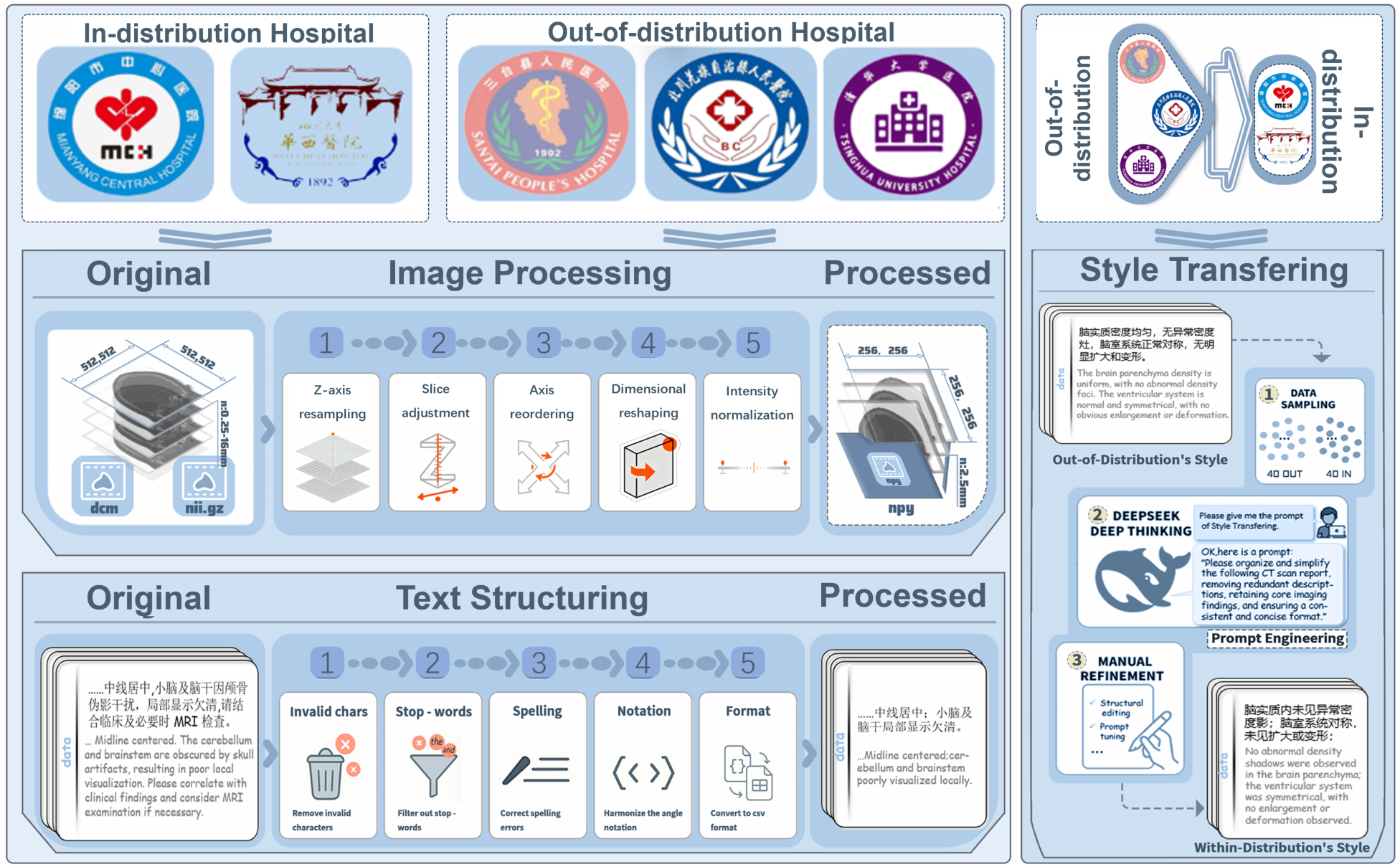
| Data sources and preprocessing for cross-hospital training and evaluation. The top panel summarizes the two in-distribution hospitals and three out-of-distribution hospitals included in the cross-hospital generalization evaluation. The middle panel illustrates the standardized image preprocessing pipeline used to convert raw CT studies into unified volumetric inputs, including z-axis resampling, slice adjustment, axis reordering, dimensional reshaping and intensity normalization. The processed volumes are saved as float32 .npy files. The bottom panel shows text structuring for Chinese radiology reports, including removal of invalid characters and normalization through stop-word filtering, spelling correction, notation harmonization and format standardization. The right panel depicts report style transfer, in which out-of-distribution reports are sampled and rewritten using a prompt-based DeepSeek workflow followed by manual refinement so that their writing style matched that of in-distribution reports while preserving clinical semantics.

Considering cross institutional differences in writing style and terminological granularity, we adopt a style transfer template based on the DeepSeek-V3 Chinese large language model to normalize expressions in out-of-distribution reports. This normalization is introduced to reduce cross-institutional heterogeneity in report formatting, expression habits, and terminology usage, which could otherwise increase unnecessary distributional variation in the text input space and impair stable multimodal learning. The LLM-based processing is restricted to structural and stylistic standardization only, with the goal of harmonizing report organization and language form across centers. It is not intended to alter, enrich, or reinterpret the original imaging findings. Original reports were retained, and the normalized reports were manually checked against the corresponding source reports during data preparation. The normalized reports were treated as harmonized report-derived references rather than independent imaging ground truth. To assess semantic preservation during report normalization, we compared prespecified clinical keyword sets between the original and normalized reports in the Santai cohort (n = 518). We calculated corpus-level micro retention recall and preservation precision, together with case-level recall, precision, Jaccard similarity, and exact keyword-set agreement (Supplementary Table 1). This analysis assessed preservation of the measured clinical keywords and did not establish equivalence of all report content.

### Emergency keyword library construction and FIES evaluation framework

To systematically quantify decision-relevant information in emergency care, we construct an emergency brain CT keyword library and, based on it, propose the FIES structured evaluation framework (Supplementary Fig. 4). The keyword library is built in a semi-automated manner. We first perform structured parsing of the original reports, then organize and consolidate emergency-related terms based on clinical expert knowledge, and further reference existing feature repositories to curate emergency-related terms that cover diverse real-world writing expressions. According to the clinical semantic hierarchy of radiology reports, keywords are categorized into three classes: imaging features, Feature, describing abnormal structural morphology, diagnostic impressions, Impression, summarizing etiology, and emergency signs, Emergency Signs, indicating critical information requiring immediate intervention. The complete bilingual keyword lexicon comprises 122 keywords across the three semantic categories (Supplementary Table 3). The construction and validation of the Impression and Emergency Signs semantic categories involved clinical expert review. Specifically, five board-certified radiologists from participating centers, each with 8–15 years of experience in neuroimaging, contributed to the definition and refinement of the semantic lexicon. To improve clinical validity and reduce annotation bias, the experts independently reviewed candidate keyword sets and their mappings to report text, followed by consensus discussions to finalize the semantic definitions and category boundaries. This hierarchical design corresponds to a complete cognitive chain from imaging findings to diagnostic conclusions and then to emergency management, thereby aligning the evaluation process closely with clinical interpretation logic.

During the matching and scoring stages of FIES, the system extracts three keyword sets from the model-generated report and the ground truth reference report, respectively, and it jointly applies exact matching and fuzzy matching mechanisms during matching to enhance robustness to lexical diversity in clinical text (Supplementary Fig. 5). For example, if the lexicon entry is “硬膜下血肿” (“subdural hematoma”) but the report uses a near-synonymous expression such as “硬膜下积血” (“subdural blood collection”), exact matching may miss the term, whereas fuzzy matching can still recover the match based on string similarity. Based on these matching results, the algorithm counts matched true positives (TP), terms appearing only in the model report (OUT), and all keywords in the reference report (GT), and it then computes precision and recall for Feature, Impression, and Emergency Signs, respectively. This structured multidimensional metric system quantifies the model’s ability to capture critical information at different semantic levels, addressing the limitation that conventional n-gram metrics such as BLEU and ROUGE are insensitive to clinical semantics in medical settings and cannot comprehensively reflect information accuracy.

### Model inputs and retrieval augmented prompting

In complex or atypical cases, a base model that relies solely on image language alignment is prone to producing safe descriptions that are factually accurate yet diagnostically shallow, in which the radiologic wording remains correct but lacks diagnostic depth and decision-oriented reasoning. To mitigate this tendency while keeping the system auditable and reproducible, we adopt a lightweight knowledge-augmented prompting strategy (Supplementary Fig. 6).

Specifically, we first construct a high-quality and structured external knowledge base by systematically curating content directly relevant to emergency brain CT interpretation from authoritative radiology reviews, clinical guidelines, and high-impact journals, and we store key imaging features, diagnostic criteria, and differential considerations for acute neurologic emergencies in a canonicalized form. For example, knowledge that “急性颅内出血在非增强 CT 上通常表现为边界清晰的高密度灶, CT 值约为 50 至 100 HU” (“acute intracranial hemorrhage on non contrast CT typically appears as a well demarcated hyperdense focus, with an attenuation of approximately 50 to 100 HU”) is encoded as a retrievable knowledge unit. This structured knowledge base is annotated under FIES semantics and uses the FIES lexicon for keyword matching, thereby combining the efficiency of sparse retrieval best matching 25 for keyword matching with the semantic understanding advantages of dense vector retrieval. When the model receives a CT case, the system first generates the corresponding case text description and maps it into the FIES semantic space as a query, and then retrieves relevant knowledge entries through hybrid retrieval. In this process, sparse retrieval is used to rapidly filter highly relevant candidates within the corresponding FIES dimension, while dense retrieval performs semantic reranking over the candidates, thereby yielding the knowledge units that best match the current imaging features and clinical semantics.

During input assembly, the retrieved background knowledge is appended to the textual prompt, “请用中文详细描述此次三维医学影像扫描的结果, 包括所有观察到的异常发现及其特征。” (“Please describe in detail in Chinese the findings of this 3D medical imaging examination, including all observed abnormalities and their characteristics.”), and, together with the visual tokens, is provided as conditional input to the language model, thereby explicitly injecting clinical priors into the generation process. Compared with the setting without retrieval augmentation, in which the prompt may only elicit radiologic descriptions, incorporating RAG enables the model to integrate imaging evidence with external knowledge and generate analysis oriented conclusions with clearer diagnostic direction and higher clinical value. For example, without retrieval augmentation, the prompt may only guide the model to generate an imaging description such as “左侧基底节区见不规则高密度灶” (“an irregular hyperdense focus in the left basal ganglia”), whereas with retrieval augmentation, the model can combine imaging features with knowledge base content to generate a more diagnostically specific conclusion, such as “左侧基底节区见不规则高密度灶……考虑急性高血压性脑出血可能大” (“an irregular hyperdense focus in the left basal ganglia … most consistent with acute hypertensive intracerebral hemorrhage”).

### Multimodal representation learning and alignment

The overall model adopts an end-to-end multimodal generative framework that jointly models 3D brain CT volumes and clinical text tasks (Fig. 9). The model consists of a 3D visual encoder, a multimodal projection module, a causal language decoder, and a parallel task output head. The overall information flow proceeds from imaging representation to semantic alignment and then to language generation. The preprocessed 3D CT volume is first encoded by the visual encoder to extract 3D representations, which is then mapped into the language model hidden space via multimodal projection. The projected representations are then fed into the language decoder as conditional inputs, together with textual prompts, enabling the model to continuously access imaging evidence during autoregressive generation. At the output stage, on top of shared multimodal semantic representations, the model executes generation and discrimination tasks in parallel. It generates a preliminary radiology report draft while simultaneously predicting emergency severity through a classification head, thereby providing both narrative reporting support and an actionable prioritization signal within a single forward pass. In the intended workflow, these outputs are used together: the urgency label supports radiology case prioritization, whereas the draft report supports rapid review, completion, and downstream communication.

**Fig. 9.**
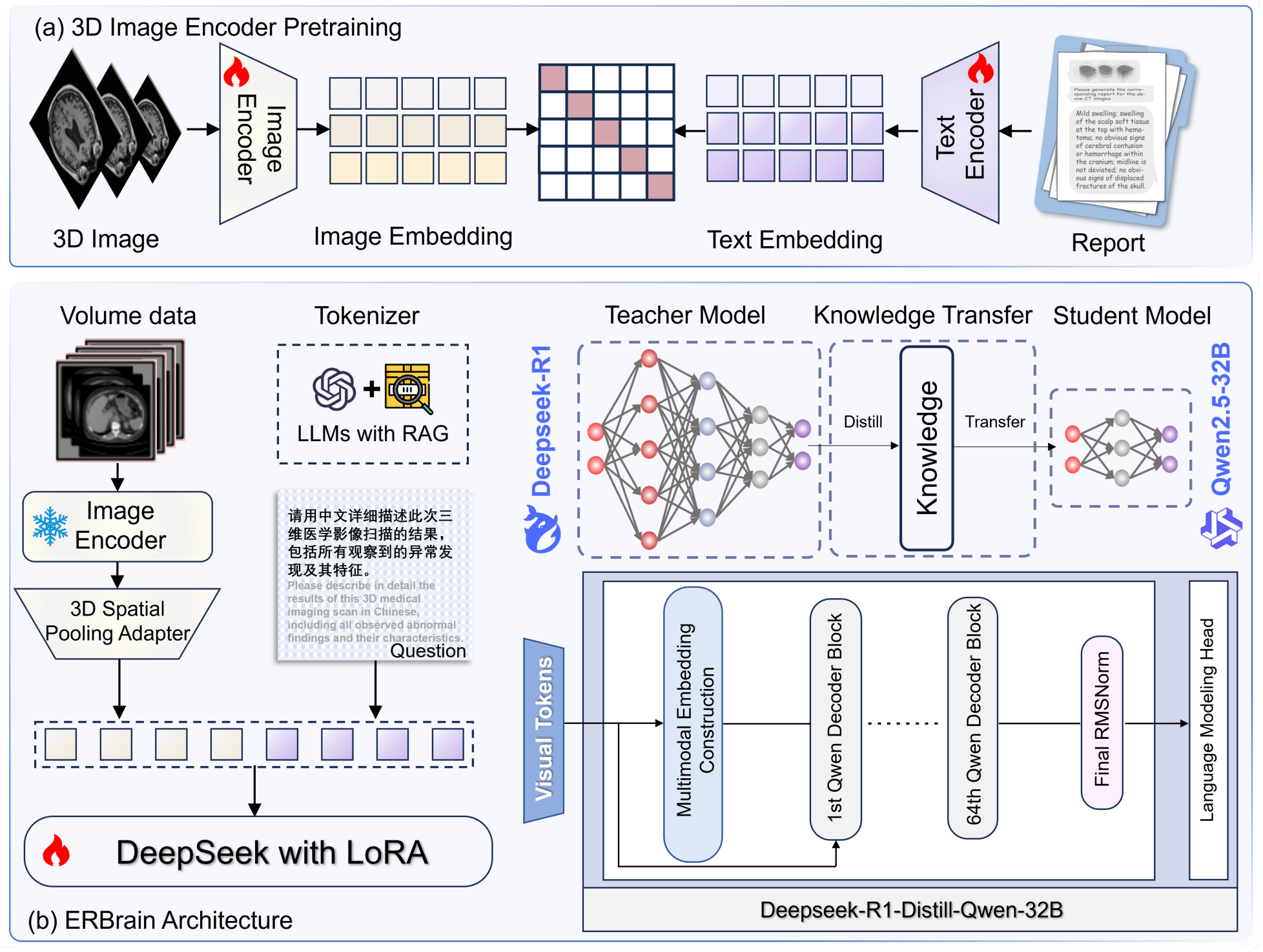
| ERBrain architecture and training initialization. a, 3D image encoder pretraining aligns volumetric image representations with report text representations by mapping the 3D image and its paired report into image and text embeddings and optimizing their cross-modal correspondence. b, ERBrain takes the preprocessed 3D volume as input, encodes it with a 3D image encoder followed by a 3D spatial pooling adapter to form compact visual tokens, and combines these tokens with an instruction prompt generated by an LLM-based tokenizer with retrieval augmentation. The multimodal tokens are then decoded by a DeepSeek-R1-Distill-Qwen-32B backbone adapted with LoRA and gated cross-attention layers inserted into the Qwen decoder blocks. The backbone is initialized with a distilled variant of DeepSeek-R1, in which reasoning capability from the original DeepSeek-R1 teacher model has been transferred to a Qwen2.5-32B student backbone to enable efficient inference.

### 3D visual encoder module

The visual backbone is a 3D Vision Transformer (3D ViT) that learns patch-level semantic representations from the preprocessed 3D brain CT volume. In emergency settings, critical lesions often manifest as small volume abnormalities distributed across slices or subtle structural shifts, and modeling based solely on 2D slices struggles to stably capture their spatial correlations. Therefore, we perform holistic modeling at the volumetric level to fully exploit 3D spatial context. In implementation, the 3D CT volume is first partitioned into 3D patches ∈ ℝ^*B×N×D_vis_*^ and then mapped into a high-dimensional token sequence, which is processed by the 3D ViT backbone via self-attention, thereby encoding both local anatomical details and global spatial structure. Through this 3D self-attention modeling, the visual encoder integrates cross-slice spatial context at the volume level and provides spatially coherent 3D visual representations for subsequent multimodal processing.

### Multimodal projection module

To reduce the high-dimensional overhead of 3D ViT outputs and suppress local noise, we introduce a 3D Spatial Pooling Perceiver (SPP) that reconstructs the patch sequence into a 3D grid and performs downsampling aggregation, preserving local global mixed semantics and making visual representations more compact and robust. The spatially aggregated visual features are further mapped by the multimodal projection module into a representation compatible with the language model hidden space ∈ ℝ^*B×N×D*_text_^, enabling visual information to participate in subsequent language decoding in the form of visual tokens.

### Causal language decoding model

After obtaining stable 3D visual representations, the model must still address the problem of accurately mapping spatial and pathologic information in images into clinical language expressions. If relying solely on autoregressive generation of the language model, the outputs are often dominated by general language priors, leading to descriptions that are inconsistent with imaging evidence. To reduce this risk of semantic drift, we introduce an explicit vision language alignment mechanism between visual encoding and language decoding so that generation is consistently constrained by imaging evidence. Specifically, the 3D features produced by the visual encoder are first mapped by the multimodal projector into a representation compatible with the language model hidden space, enabling visual information to participate in subsequent autoregressive decoding as visual tokens. By jointly modeling visual and textual features in a unified semantic space, the model can dynamically leverage imaging cues during generation, thereby reducing language outputs that deviate from imaging evidence. On this basis, the model adopts a causal language modeling paradigm and performs token-by-token autoregressive generation of radiologic descriptions under visual conditioning in a unified manner to support the expression of complex emergency clinical semantics and diagnostic reasoning.

### Parallel task output head

For language modeling, we adopt a causal language model from the Qwen family and initialize it with the publicly available distilled checkpoint DeepSeek-R1-Distill-Qwen-32B. Knowledge distillation is a model compression paradigm in which the reasoning behavior of a large teacher model is transferred to a smaller or more efficient student model, thereby retaining much of the teacher’s reasoning capability while reducing inference-time computational cost. In our configuration, DeepSeek-R1 serves as the teacher whose reasoning competence is encoded in the distilled parameters, whereas Qwen2.5-32B acts as the student backbone, offering efficient Chinese generation and deployment-friendly characteristics. This teacher–student distillation scheme preserves the deep reasoning capacity of DeepSeek-R1 while leveraging Qwen’s general language modeling strengths, forming the core engine for system-level generation and comprehension.

Given the prohibitive computational and data requirements of full-parameter fine-tuning for large language models, we employ a parameter-efficient fine-tuning (PEFT) strategy for multimodal adaptation. This design allows the model to maintain its pretrained linguistic competence while acquiring image-conditioned generation capability tailored to emergency brain CT. The multimodal adaptation stage primarily optimizes vision–language alignment, establishing a stable cross-modal semantic space that supports subsequent emergency-oriented training constraints. Building upon the shared multimodal representation, we further introduce a parallel output-head design: in addition to generating complete radiology reports, the model simultaneously performs structured discriminative tasks relevant to emergency settings. As a result, natural-language descriptions and structured decision signals can be produced within a single forward pass.

### Joint training objectives and emergency priority modeling

In emergency brain CT scenarios, model training must not only generate radiology reports that are linguistically fluent and semantically coherent, but also remain highly sensitive to clinically urgent signs while minimizing language hallucinations that deviate from imaging evidence. To this end, we construct a joint training objective jointly constrained by generation quality, cross-modal consistency, and emergency prioritization. The overall loss function comprises four components: an autoregressive generation loss, a vision-language alignment loss, an emergency keyword weighted loss, and a classification cross-entropy loss that is included when structured labels are available (Supplementary Fig. 7).

### Autoregressive generation loss

The autoregressive generation loss adopts the standard autoregressive cross-entropy for causal language models. Let the logits predicted by the model at position *t* for sample *b* be *z_b,t_* ∈ 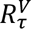, and let the ground truth label be *y_b,t_* (positions to be ignored are marked as -100). The generation loss is then defined as:

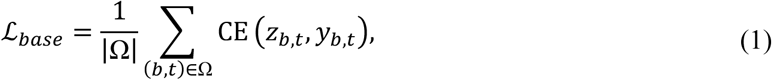

where Ω denotes the set of all valid token positions. This term is implemented within the HuggingFace framework and is computed only when text labels are provided.

### Vision-language alignment loss (DTrace Loss)

To reduce excessive reliance on language priors during generation, we introduce a vision-language alignment loss, DTrace Loss, to constrain consistency between linguistic semantics and global visual representations. Given the visual projected feature sequence for sample 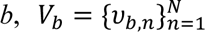, and the decoder’s last position language feature **h***_b_*, we define the mean visual vector 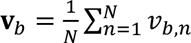, and adopt an alignment loss in the form of mean squared error:

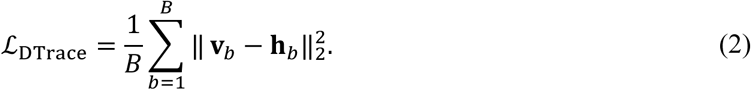

This term is incorporated into the total loss with a weighting coefficient *λ_d_* (default *λ_d_* = 1.0) to encourage language generation to rely on visual evidence at the global semantic level.

### Emergency keyword weighted loss (Critical Loss)

In clinical emergency settings, a small set of phrases often directly determines treatment priority, such as “急性颅内出血” (“acute intracranial hemorrhage”) and “中线移位大于 5 mm” (“midline shift > 5 mm”). To explicitly amplify the importance of these phrases during training, we design Critical Loss by assigning higher weights to token positions corresponding to critical phrases contained in the reference labels, so that prediction errors at these positions are magnified and preferentially learned. Specifically, based on a preconstructed emergency keyword library, we locate the corresponding phrases within the reference text and map them to token-level positions; matched positions are assigned weight *w_emg_*, whereas all other positions use the base weight *w_base_*. Let the per token unreduced cross entropy be *E_b,t_*, the valid mask be *m_b,t_*, and the weight matrix be *W_b,t_*. Then,

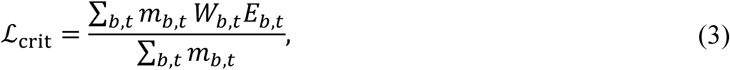

where

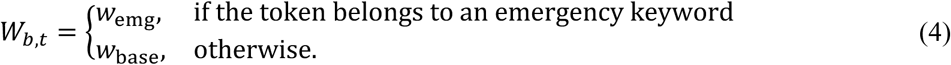

By default, we set *w*_emg_=2.0,*w*_base_=1.0. This design preserves the overall loss scale while concentrating learning signals on expressions that are decisive for clinical decision making.

### Classification loss

When a sample includes a case level structured label *c_b_* ∈ {1, …, *K*}, the model outputs logits *s_b_* through a parallel classification head and adopts the standard cross entropy loss:

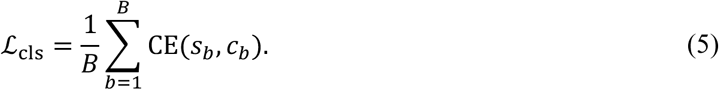

### Overall loss function

During training, all loss terms are combined with empirically chosen coefficients:

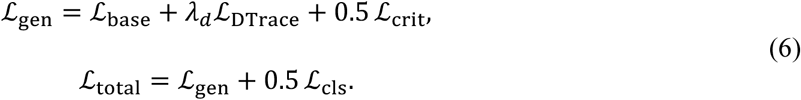

The coefficients for Critical Loss and the classification loss are set to 0.5. If the corresponding labels are not provided, the associated term is excluded from optimization.

### Metrics for report generation

BLEU is a widely used metric for assessing the quality of machine-generated text. It is fundamentally based on n-gram precision and measures the degree of overlap between a candidate text and a reference text across different n-gram orders. BLEU-1 to BLEU-4 correspond to 1-gram through 4-gram matches, progressively characterizing contextual consistency from the word level to the phrase level. To prevent artificially inflated scores when the candidate text is overly short, BLEU introduces a brevity penalty (BP). It is defined as:

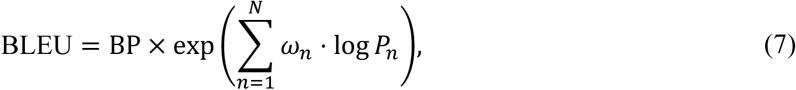

where BP denotes the brevity penalty, *P_n_* is the *n* th order n gram precision, *n* ∈ {1,2,3,4} corresponds to BLEU-1 to BLEU-4, and *ω_n_* is the weight assigned to each order.

ROUGE-L (Longest Common Subsequence) differs from n-gram-based metrics in that it quantifies overlap via the longest common subsequence (LCS) between the candidate and reference texts, thereby accounting for both word occurrence and word order. This property makes it more suitable for capturing fluency and relevance in text generation. Its F form is defined as:

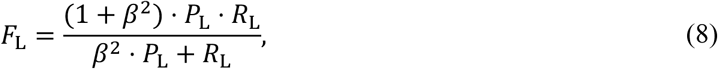

where 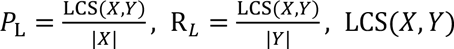 denotes the length of the longest common subsequence between the candidate text *X* and the reference text *Y*, and β is typically set to 1 to balance the relative importance of precision and recall.

BERT-Score is a semantic evaluation metric built upon pretrained contextual embeddings. It measures semantic consistency by computing cosine similarity between token representations from the candidate and reference texts, and thus can better capture semantic equivalence even when surface forms differ. The final score is reported in F form:

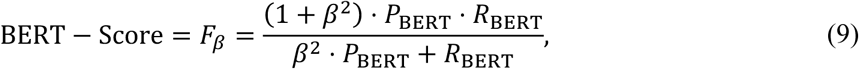

where 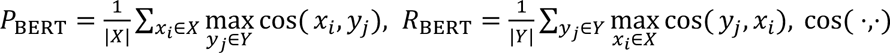 denotes cosine similarity, and β is typically set to 1 to balance precision and recall. To mitigate embedding space bias induced by different pretrained models, a baseline rescaling can be applied to normalize the raw scores:

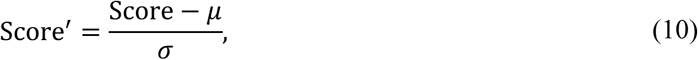

where *μ* and σ are the corresponding mean and standard deviation of the score distribution computed on the reference corpus.

METEOR establishes token-level alignments between the candidate and reference texts, including exact matches, synonym matches, and stemming matches, and computes a harmonic mean of precision and recall. It further introduces a fragmentation penalty to discourage discontinuous matches that may appear similar on the surface yet exhibit disordered word sequences. It is defined as:

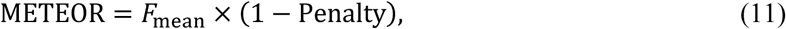

where 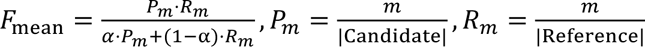, *m* is the number of matched tokens and α controls the relative weighting between precision and recall. respectively. The penalty term accounts for discontinuous matches:

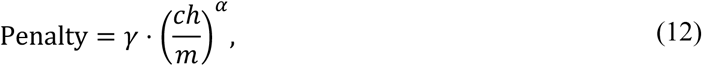

where *ch* is the number of contiguous matched segments, and γ and α are hyperparameters controlling the strength of the penalty.

### FIES clinical semantic evaluation framework

To address the limitation of generic text metrics in medical settings, which often emphasize surface matching while underweighting clinically critical information, we introduce the FIES structured clinical semantic evaluation framework (Supplementary Fig. 8). By extracting medical keywords and grouping them by semantic category, FIES quantitatively evaluates the accuracy and coverage of key clinical information in generated reports. Within this framework, a report is decomposed into three semantic dimensions that directly relate to emergency clinical decision making.

FIES-Feature evaluates the accuracy of key information generation at the imaging feature level, namely, how many imaging feature statements in the generated report are supported by the reference report. By quantifying keyword matches between imaging feature terms extracted from the generated and reference texts, this metric characterizes the reliability of the model’s expression of imaging findings. Specifically, for a given sample, let 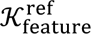 denote the set of imaging feature keywords identified from the reference report and 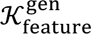 denote the set identified from the generated report. When 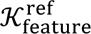 is non-empty, FIES-Feature is defined as:

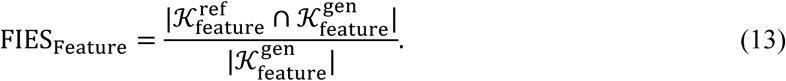

If no imaging feature keywords are identified in the generated report, the metric value is set to 0. If the reference report contains no keywords in this dimension, the sample is excluded from aggregation.

FIES-Impression evaluates the accuracy of generated content at the diagnostic impression level, namely, how many diagnosis related keywords produced by the model are consistent with the impressions in the reference report. This metric reflects the reliability of the model’s summarizing diagnostic statements without depending on the specific linguistic realization. For a given sample, let 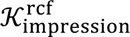 denote the set of diagnostic impression keywords identified from the reference report and 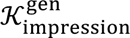 denote the set identified from the generated report. When 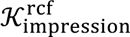 is non-empty, FIES-Impression is defined as:

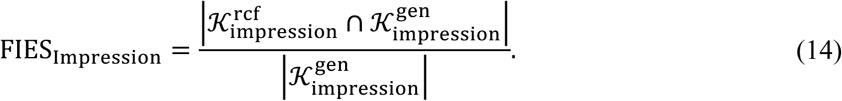

If no diagnostic impression keywords are identified in the generated report, the metric value is set to 0. If the reference report contains no keywords in this dimension, the sample is excluded from aggregation.

FIES-Emergency Signs evaluates the accuracy of key information generation at the emergency sign level, namely, how many risk related keywords in the generated report can be substantiated by corresponding evidence in the reference report. This metric focuses on whether the model reliably expresses information that is tightly coupled with clinical management. For a given sample, let 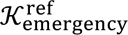 denote the set of emergency related keywords identified from the reference report and 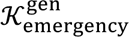 denote the set identified from the generated report. When 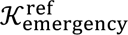 is non-empty, FIES-Emergency Signs is defined as:

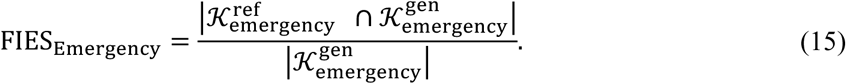

If no emergency related keywords are identified in the generated report, the metric value is set to 0. If the reference report contains no keywords in this dimension, the sample is excluded from aggregation.

### Evaluation metrics for the classification task

Macro-Precision first computes class wise precision and then takes the arithmetic mean across classes, thereby measuring overall predictive reliability across classes without being influenced by class frequency. For class *c*, precision is defined as:

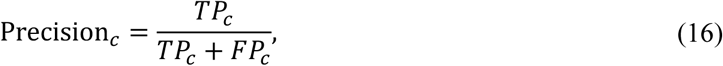

where *TP_c_* is the number of samples correctly predicted as class *c* and *FP_c_* is the number of samples incorrectly predicted as class *c*. Macro Precision is defined as:

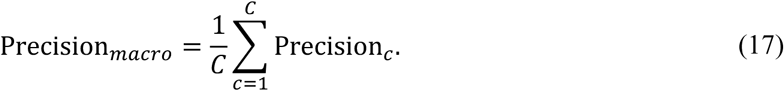

Macro Recall averages class wise recall to measure overall detection ability across classes. For class *c*, recall is defined as:

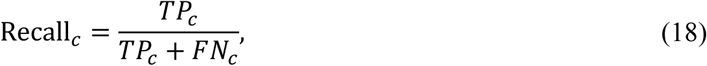

where *FN*_c_ denotes the number of samples of class *c* that are incorrectly predicted as other classes. Macro Recall is defined as:

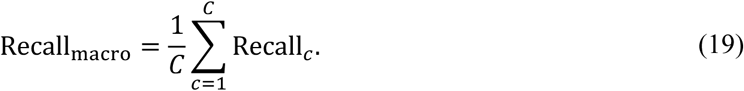

Macro-F1 is the harmonic mean of Macro Precision and Macro-Recall, jointly balancing predictive accuracy and coverage across classes. It is defined as:

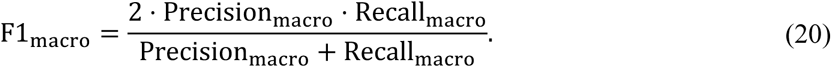

Accuracy (ACC) measures the proportion of all samples that are correctly predicted, including both positive and negative instances. It is defined as:

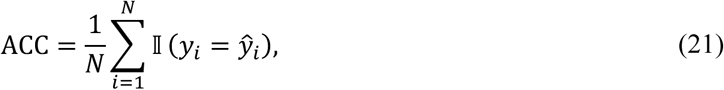

where N denotes the total number of samples, *y_i_* and *ŷ_i_* denote the ground truth and predicted label for the *i* th sample, respectively, and I is the indicator function.

Balanced Accuracy (BACC) is designed for imbalanced datasets and is computed as the average recall over all classes, thereby better reflecting per class performance. In multiclass settings, BACC is defined as:

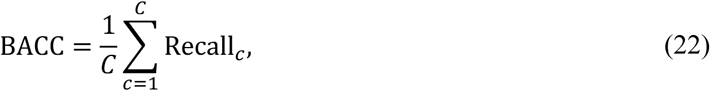

where *C* denotes the number of classes and Recall_7_ denotes recall for class *c*.

Specificity measures the model’s ability to correctly identify negative instances, namely, the proportion of true negatives that are correctly predicted as negative. This metric reflects the model’s ability to avoid false alarms and is particularly important in clinical settings with class imbalance or high costs associated with false positives. For class *c*, specificity is defined as:

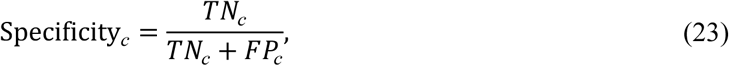

where *TN_c_* denotes the number of samples not belonging to class *c* that are correctly predicted as not class *c*, and *FP_c_* denotes the number of samples not belonging to class *c* that are incorrectly predicted as class *c*. In multiclass settings, we compute each class and then take the macro average as the overall specificity:

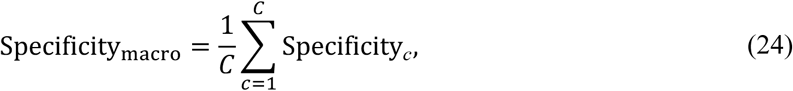

where *C* denotes the number of classes.

G-mean is an evaluation metric that jointly considers the model’s capability to recognize positives and negatives. By taking the geometric mean of sensitivity and specificity, it reflects the overall balance of performance across classes. Compared with single metrics, G-mean is more sensitive to class imbalance and can effectively characterize whether the model simultaneously achieves positive detection and negative exclusion. For class *c*, G-mean is defined as:

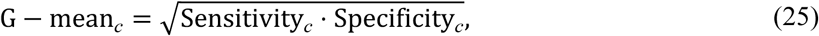

where Sensitivity*_c_* = 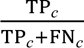, corresponds to the recall of class *c*. In multiclass settings, we likewise compute class wise G-mean values and take their macro average to obtain the overall G-mean.

### LLM Inference and Reproducibility Settings

ERBrain adopts a LLaVA-style multimodal architecture integrating a 3D visual encoder with a causal language model backbone. During inference, report generation is performed using greedy decoding without stochastic sampling. The maximum output length is set to 512 tokens, and a fixed random seed of 42 is used to ensure reproducibility. No temperature scaling, top-p sampling, or beam search strategies are applied. All dataset splits are performed at the patient level to prevent information leakage across training, validation, and testing sets. External validation centers are fully independent and not involved in model training, hyperparameter selection, or prompt modification. The FIES keyword lexicon is constructed exclusively from the training data and is frozen prior to evaluation to ensure strict separation between model development and testing.

## Ethics statement

This study receives institutional review board approval from all participating centers. Data collection at West China Hospital is approved by its Research Ethics Committee (2025-1612). Data collection at Mianyang Central Hospital is approved by its Research Ethics Committee (S202503234-1). The Research Ethics Committee of Santai County People’s Hospital approved data collection under protocol 2025-KY-0076, while Beichuan Qiang Autonomous County People’s Hospital approved data collection under protocol LL-20250002. In addition, data collection at Tsinghua University Hospital is approved by its Research Ethics Committee (THU-01-2025-1054-R1). All patient data are anonymized and de-identified prior to analysis. Given the retrospective nature of the study, the requirement for informed consent is waived. All procedures are conducted in accordance with the Declaration of Helsinki and relevant ethical guidelines for medical research.

## Data availability

The emergency brain CT images, associated Chinese radiology reports, and derived annotations used in this study are collected retrospectively from multiple clinical centers and contain sensitive patient information. Therefore, these data are not publicly available due to patient privacy protection regulations and institutional data governance policies. Researchers seeking access to the data for legitimate scientific and non-commercial research purposes may submit a reasonable request to the corresponding author. All requests will be evaluated on a case-by-case basis and are subject to approval by the Ethics Committees of the participating institutions, including West China Hospital, Mianyang Central Hospital, Santai County People’s Hospital, Beichuan Qiang Autonomous County People’s Hospital, and Tsinghua University Hospital. The Ethics Committees reserve the right to approve or deny data access requests.

## Code availability

The complete source code for model construction and implementation, including data preprocessing pipelines, model architecture, and evaluation scripts, is available for download through GitHub (https://github.com/JustlfC03/ERBrain). This code repository contains detailed documentation and usage instructions to facilitate reproduction of our results and further development of the models.

## Acknowledgments

This work is supported by the National Natural Science Foundation of China (grant number 82302166), Dushi Program (grant number 20261080018), Tsinghua University Startup Fund, and Natural Science Foundation of Sichuan Science and Technology Department (2024NSFSC0656).

## CRediT authorship contribution statement

Yifei Chen: Methodology, Conceptualization, Writing – original draft, Writing – review & editing. Jingyuan Zheng: Methodology, Writing – original draft. Yuanhan Wang: Methodology, Image-to-text experiments. Beining Wu: Image-to-text experiments, Visualization. Lu Li: Data collection (West China Hospital), Annotation. Mingxuan Liu: Image-to-text experiments. Liaoman Xu: Classification experiments. Yueyi Wu: Classification experiments. Chang Liu: Visualization. Lulu Guo: Visualization. Hongjia Yang: Writing – review & editing, Xuguang Bai: Classification experiments, Feiwei Qin: Writing – review & editing, Qiang Liao: Data collection (Beichuan Qiang Autonomous County People’s Hospital), Annotation, Yong Gu: Data collection (Santai County People’s Hospital), Annotation, Gang Zhao: Data collection (Santai County People’s Hospital), Annotation, Lu Ma: Data collection (Tsinghua University Hospital), Annotation, Keqin Pan: Data collection (Tsinghua University Hospital), Annotation, Jun Guo: Data collection (Tsinghua University Hospital), Annotation, Ying Zhou: Data collection (Mianyang Central Hospital), Annotation, Huaiqiang Sun: Supervision, Writing – review & editing, Qiyuan Tian: Methodology, Conceptualization, Supervision, Project administration, Funding Acquisition, Resources, Writing – original draft, Writing – review & editing.

