## Supplementary Information for "A retrospective study of a Chinese vision-language large model for emergency 3D brain CT interpretation"

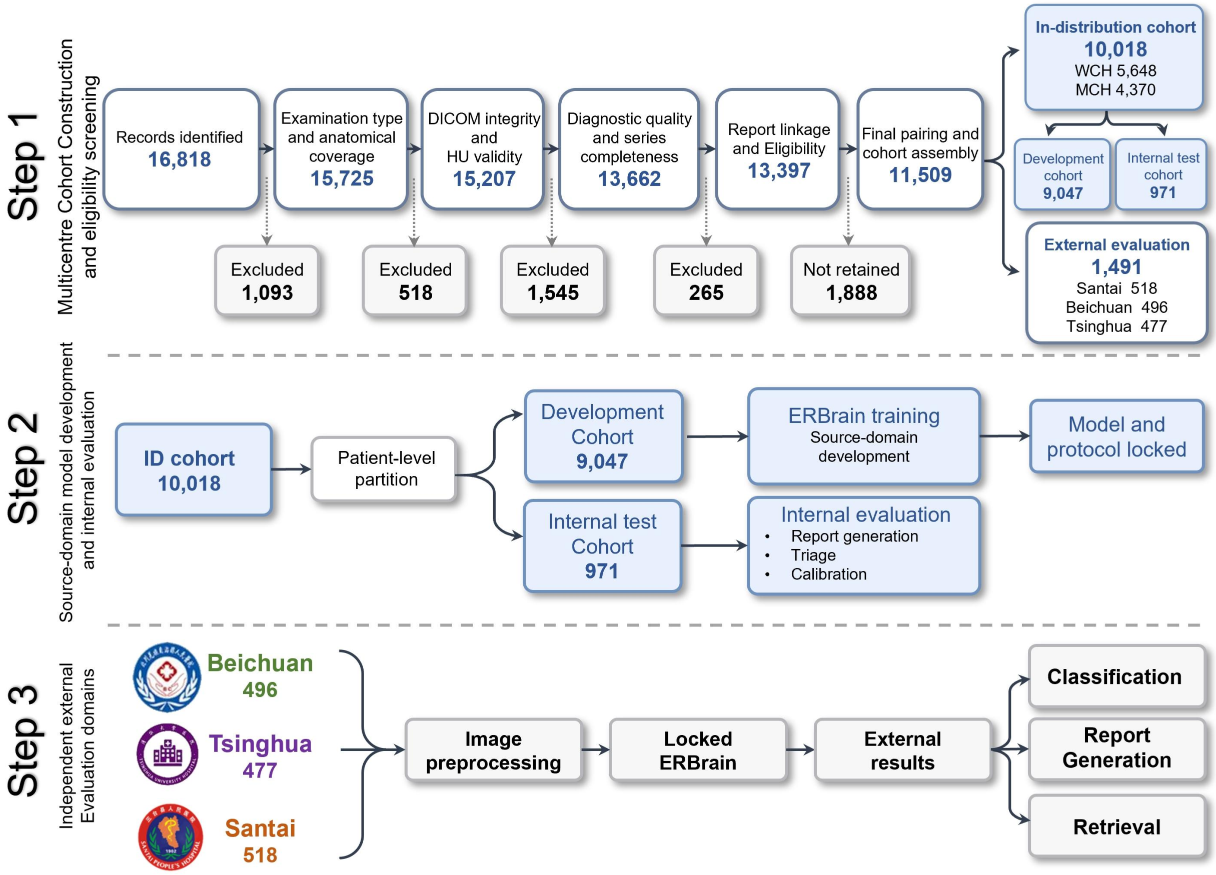

**Supplementary Fig. 1 | Multicenter cohort construction, data partitioning, and evaluation design.** The schematic summarizes the three-stage design of the study. In Step 1, multicenter cohort construction and eligibility screening reduced 16,818 identified records to 11,509 paired studies through successive filtering on examination type and anatomical coverage, DICOM integrity and HU validity, diagnostic quality and series completeness, and report linkage and eligibility; the in-distribution cohort (n = 10,018; West China Hospital 5,648 and Mianyang Central Hospital 4,370) was partitioned into a development cohort (n = 9,047) and an internal test cohort (n = 971), and three additional hospitals (Santai 518, Beichuan 496, and Tsinghua 477; n = 1,491) were held out for external evaluation. In Step 2, the development cohort was used for ERBrain training and the internal test cohort for report generation, triage, and calibration, after which the model and evaluation protocol were locked. In Step 3, the locked ERBrain was applied to the three external out-of-distribution domains after unified image preprocessing to assess classification, report generation, and retrieval.

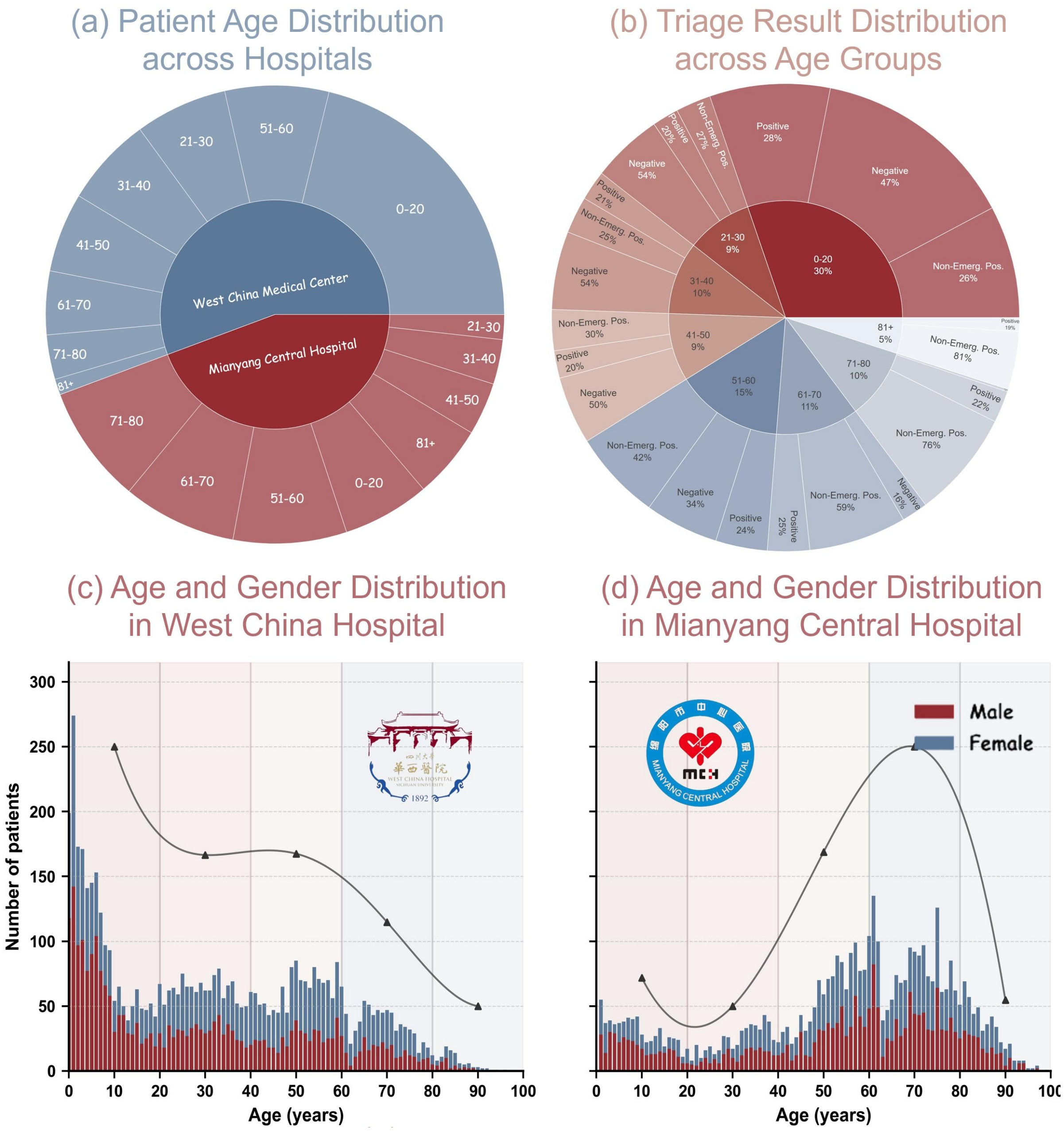

**Supplementary Fig. 2 | Demographic characteristics and triage label distributions of the in-distribution cohorts, and multicenter data inclusion.** a, Patient age distribution across hospitals, with age grouped into predefined bins. b, Distribution of emergency severity triage outcomes across age groups, showing the proportions of Negative, Non-emergency-positive, and Positive cases within each age bin. c, Age and gender distribution in West China Hospital. d, Age and gender distribution in Mianyang Central Hospital.

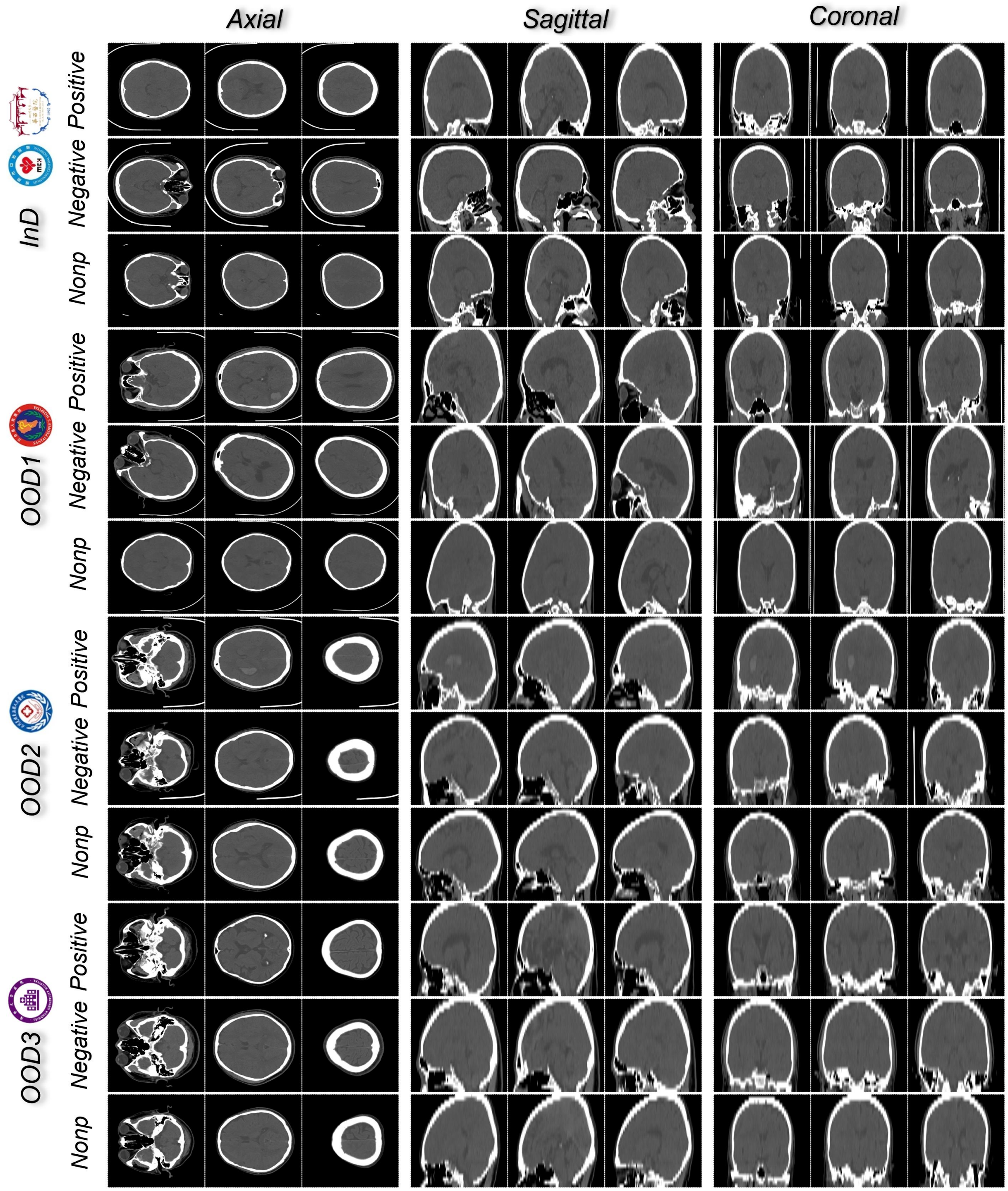

**Supplementary Fig. 3 | Representative emergency brain CT examples across cohorts and triage categories.** Each row shows a representative case from the in-distribution cohort (InD) or one of the three out-of-distribution cohorts (OOD1-OOD3), stratified by triage category into Positive, Negative, and Non-emergency-positive (Nonp). For every case, axial, sagittal, and coronal reconstructions are displayed to illustrate cross-center differences in acquisition protocol, slice profile, and image appearance.

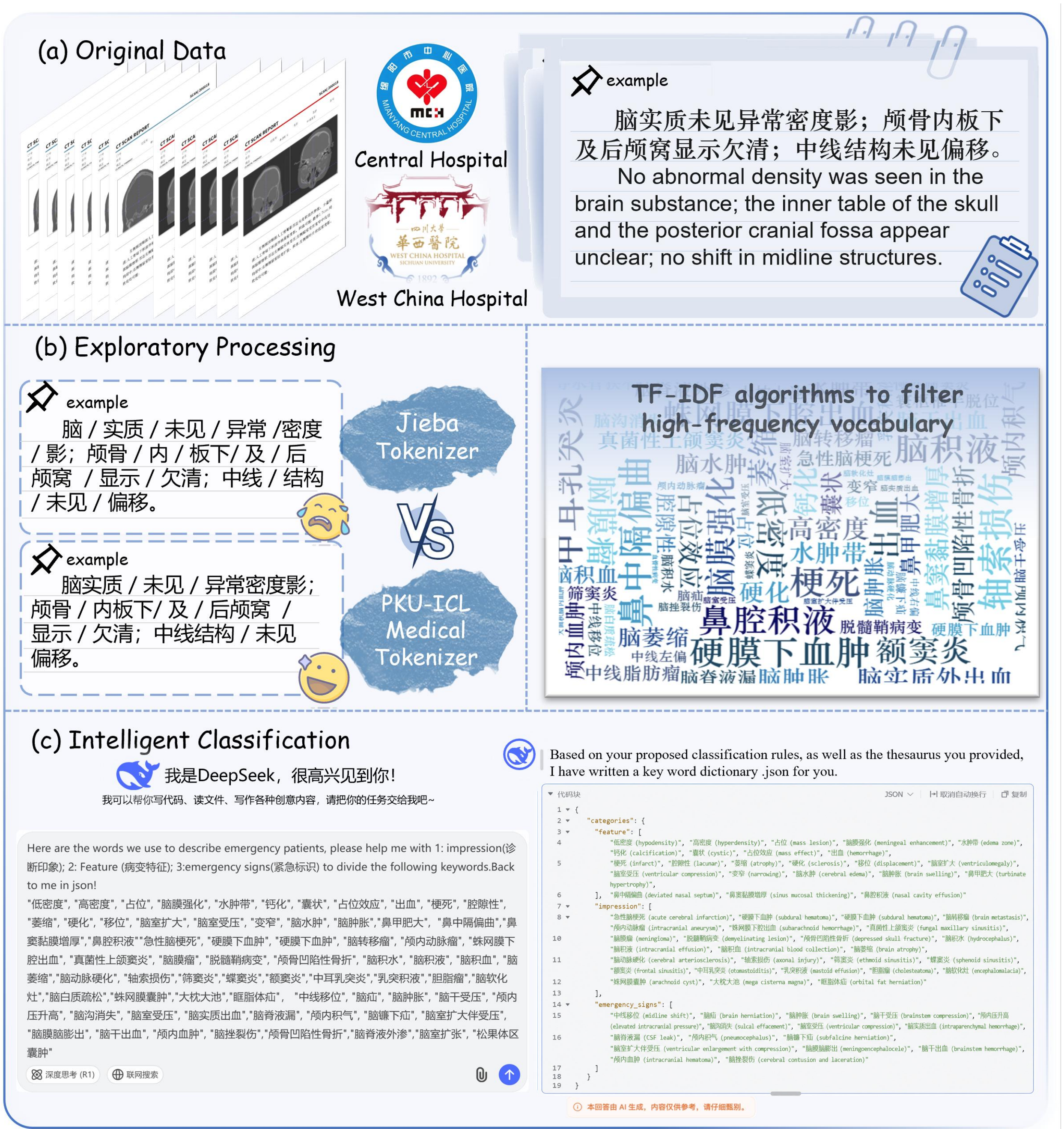

**Supplementary Fig. 4 | Construction of the emergency brain CT keyword library.** a, Original radiology reports collected from the participating centers. b, Exploratory processing, in which a domain-adapted medical segmenter is compared against a general-purpose word segmenter and high-frequency clinical terms are extracted using TF-IDF weighting. c, Intelligent classification, in which candidate terms are organized with large-language-model assistance and expert review into the Feature, Impression, and Emergency Signs categories, yielding the structured keyword dictionary that underlies the FIES framework.

**Algorithm: FIES Evaluation Framework**

**Input**: Generated Text $G$, Reference Text $R$, Keyword Library $K$, Threshold $T$

**Output**: Precision $P$, Recall $R$, True Positive Count $TP$

Initialize keyword library $K$ from configuration file
Build keyword set $K_{set}$ and category map $K_{map}$ for fast lookup
Define fuzzy matching function $F(text, dim, K, T)$:
    Initialize empty set $found$
    For each keyword $kw$ in $K_{map}[dim]$:
        If $kw$ in $text$:
            Add $kw$ to $found$
    Tokenize $text$ using $jieba$
    For each keyword $text$ in $K_{map}[dim]$:
        If $kw$ not in $found$:
            Use $get_{close_{matches}}(kw, tokens, n=1, cutoff=T)$
            If match found:
                Add $kw$ to $found$
    Return $found$
For each sample $(G, R)$ in dataset:
    For each dimension $dim$ in $dims$:
        $GT_{terms} \leftarrow F(R, dim, K, T)$
        $OUT_{terms} \leftarrow F(G, dim, K, T)$
        $TP \leftarrow |GT_{terms} \cap OUT_{terms}|$
        If $|OUT_{terms}| = 0$:
            $Precision \leftarrow 0$
        Else:
            $Precision \leftarrow \frac{TP}{|OUT_{terms}|}$
        $Recall \leftarrow \frac{TP}{|GT_{terms}|}$
        Append $P$ and $R$ to respective lists
Return
    Per-dimension average $P$ and $R$
    Overall average $P$ and $R$

**Supplementary Fig. 5 | Pseudocode for the FIES evaluation framework.** Given a generated report *G*, a reference report *R*, a keyword library *K*, and a fuzzy-matching threshold *T*, the algorithm first initializes a per-dimension keyword map for fast lookup, and then extracts dimension-specific keyword sets from *G* and *R* using exact matching together with token-based fuzzy matching. For each semantic dimension, matched terms are counted as true positives (*TP*), and precision and recall are computed from the extracted sets. The algorithm outputs per-dimension precision and recall, and it further aggregates them to obtain overall averaged scores across dimensions and across samples.

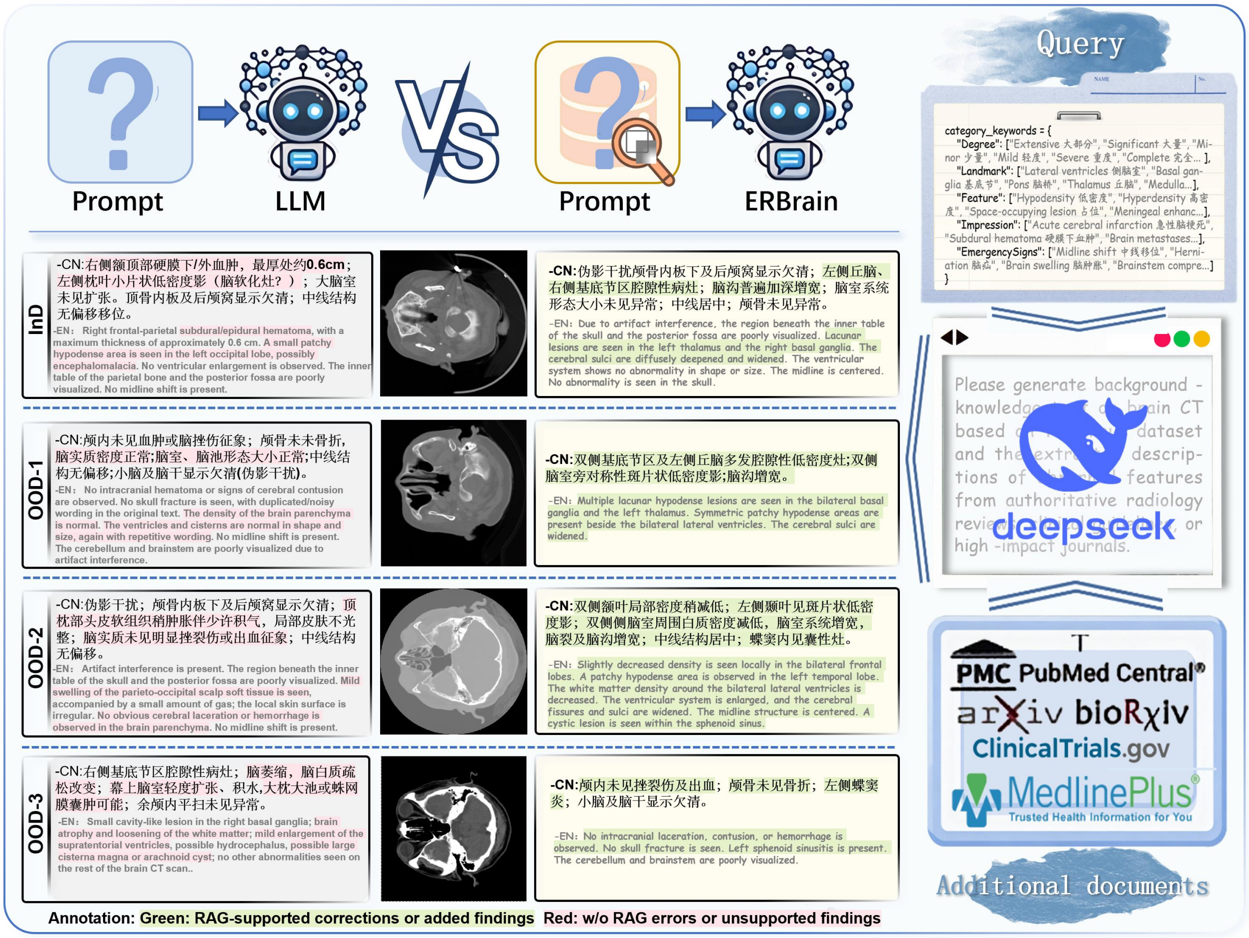

**Supplementary Fig. 6 | Lightweight knowledge-augmented prompting strategy for report generation.** Reports generated by a general large language model and by ERBrain are compared across the in-distribution and three out-of-distribution cohorts. Category-specific keywords are used to retrieve background knowledge from authoritative biomedical sources, which is supplied as auxiliary context during generation. Green text denotes retrieval-supported corrections or added findings, whereas red text denotes errors or unsupported findings produced without retrieval augmentation.

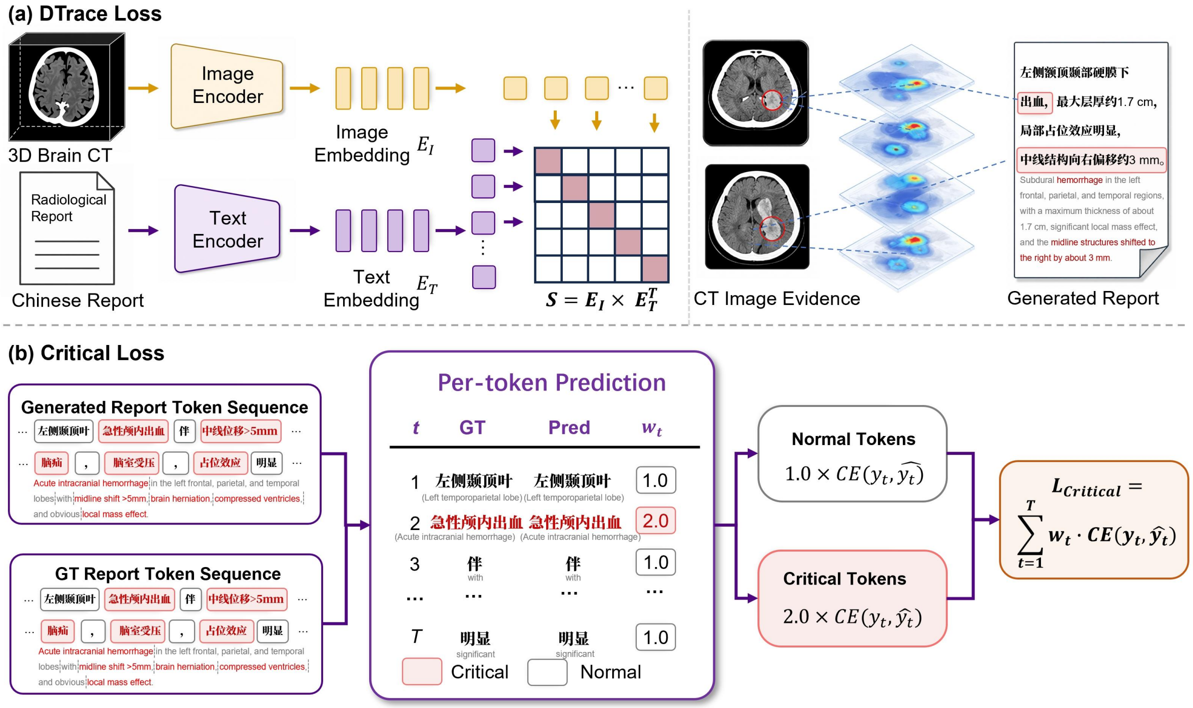

**Supplementary Fig. 7 | Joint training objective of ERBrain.** a, DTrace loss aligns paired brain CT volumes and radiology reports: a 3D image encoder and a text encoder project them into a shared embedding space, and the resulting similarity matrix is optimized so that imaging evidence is matched to the corresponding report content. b, Critical loss reweights the per-token generation objective by clinical importance, so that emergency-critical tokens contribute more strongly than routine tokens to the overall loss.

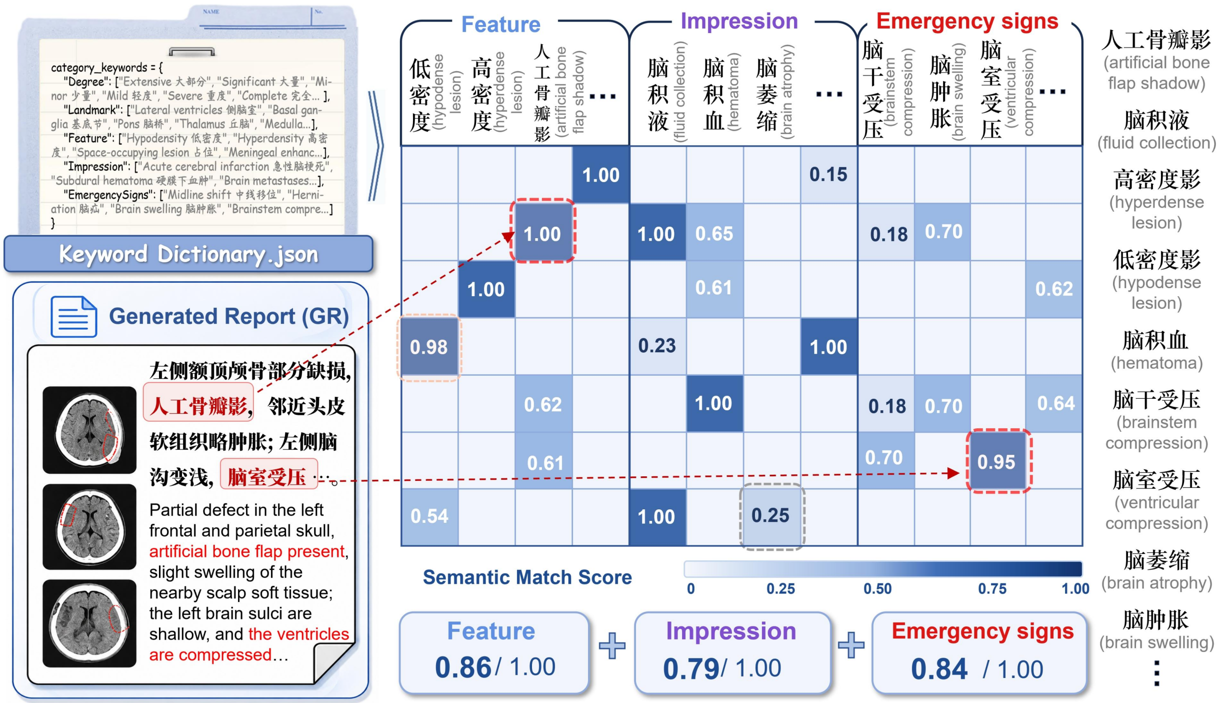

**Supplementary Fig. 8 | FIES structured clinical semantic evaluation framework.** Keywords extracted from a generated report are matched against the reference lexicon within three semantic dimensions (Feature, Impression, and Emergency Signs) using combined exact and fuzzy matching. The resulting semantic match-score matrix quantifies agreement for each dimension, and the per-dimension scores are aggregated into an overall FIES assessment of the clinically relevant information contained in the report.

| Metric | All OOD (n=1,491) | Santai (n=518) | Beichuan (n=496) | Tsinghua (n=477) |
|---|---|---|---|---|
| **Report-level concordance** | | | | |
| Original keyword-positive reports | 739 (49.6%) | 280 (54.1%) | 296 (59.7%) | 163 (34.2%) |
| Normalized keyword-positive reports | 684 (45.9%) | 269 (51.9%) | 256 (51.6%) | 159 (33.3%) |
| Keyword-positive in both versions | 665/1491 (44.6%); 665/739 (90.0%) | 262/518 (50.6%); 262/280 (93.6%) | 244/496 (49.2%); 244/296 (82.4%) | 159/477 (33.3%); 159/163 (97.5%) |
| Original-positive only | 74 (5.0%) | 18 (3.5%) | 52 (10.5%) | 4 (0.8%) |
| Normalized-positive only | 19 (1.3%) | 7 (1.4%) | 12 (2.4%) | 0 (0.0%) |
| Keyword-negative in both versions | 733 (49.2%) | 231 (44.6%) | 188 (37.9%) | 314 (65.8%) |
| **Occurrence-level preservation** | | | | |
| Original keyword occurrences | 1,479 | 717 | 536 | 226 |
| Normalized keyword occurrences | 1,218 | 634 | 370 | 214 |
| Retained original keyword occurrences | 1,155 | 610 | 331 | 214 |
| Dropped original keyword occurrences | 324/1479 (21.9%) | 107/717 (14.9%) | 205/536 (38.2%) | 12/226 (5.3%) |
| Newly introduced keyword occurrences | 63/1218 (5.2%) | 24/634 (3.8%) | 39/370 (10.5%) | 0/214 (0.0%) |
| Micro keyword retention recall | 78.1% | 85.1% | 61.8% | 94.7% |
| Micro keyword preservation precision | 94.8% | 96.2% | 89.5% | 100.0% |
| Net occurrence change | -261/1,479 (-17.6%) | -83/717 (-11.6%) | -166/536 (-31.0%) | -12/226 (-5.3%) |
| **Case-level keyword-set agreement** | | | | |
| Mean case-level retention recall | 79.1% | 84.7% | 64.8% | 95.6% |
| Mean case-level preservation precision | 93.4% | 94.9% | 87.8% | 100.0% |
| Mean case-level Jaccard similarity | 76.0% | 81.3% | 60.8% | 95.6% |
| Exact keyword-set agreement | 1,219 (81.8%) | 422 (81.5%) | 331 (66.7%) | 466 (97.7%) |
| Non-identical keyword sets | 272 (18.2%) | 96 (18.5%) | 165 (33.3%) | 11 (2.3%) |

**Supplementary Table 1 | Preservation of clinical keywords after DeepSeek-V3 report normalization.** The analysis included 518 paired original and normalized reports from Santai County People's Hospital. Corpus-level metrics quantify the retention of clinical keyword occurrences across the complete cohort, whereas case-level metrics compare the keyword sets extracted from each original report and its normalized counterpart. Exact keyword-set agreement indicates that identical clinical keyword sets were identified in the two report versions. These results assess preservation of the measured clinical keywords and do not establish equivalence of all report content.

| Parameter | In Distribution | | Out of Distribution | | |
|---|---|---|---|---|---|
| | Center1 | Center2 | Center3 | Center4 | Center5 |
| DICOM Records | 140513 | 5,648 | 12752 | 199063 | 29362 |
| Patient Records | 4135 | 5,407 | 518 | 496 | 477 |
| Male(%) | 2,062 (49.9%) | 2,660 (49.2%) | 254 (49.0%) | 207 (41.7%) | 232 (48.6%) |
| Female(%) | 2,073 (50.1%) | 2,747 (50.8%) | 264 (51.0%) | 289 (58.3%) | 247 (51.4%) |
| Age(Avg) | 53 | 30.7 | 63 | 54 | 47 |
| CT tube Voltage | 100~120KV | 80-150KV | 120KV | 100~140KV | 140KV |
| CT tube Current | 288.8 (30-600) | 285 (10-380) | 295 (10-380) | 155 (39-464) | 62.8 (24-506) |
| Slice Thickness | 5.0mm | 2.5mm | 2.5mm | 1.25mm | 5.0mm |
| CTScanner | GE | GE (41.7%)<br>Siemens(58.52%) | GE (29.7%)<br>Philips (52.6%)<br>UIH (17.7%) | UIH | Siemens |

**Supplementary Table 2 | Scanner configurations and acquisition parameters across the five participating centers.** For each center, the table summarizes the number of DICOM and patient records, sex and age composition, and representative CT acquisition settings, including tube voltage, tube current, slice thickness, and scanner manufacturer. Centers are grouped into in-distribution and out-of-distribution sources to document the cross-institutional differences in acquisition hardware and protocol that underlie the distribution shifts evaluated in this study.

**Bilingual FIES Keyword Lexicon**

```
{
  "feature": [
    "低密度(Low density)，高密度(High density)，占位(Space-occupying lesion)，脑膜强化(Meningeal enhancement)，
水肿带(Edema zone)，钙化(Calcification)，囊状(Cystic)，占位效应(Mass effect)，出血(Hemorrhage)，梗死(Infarction)，
腔隙性(Lacunar)，萎缩(Atrophy)，硬化(Sclerosis)，移位(Displacement)，脑室扩大(Ventricular enlargement)，
脑室受压(Ventricular compression)，变窄(Narrowing)，脑水肿(Cerebral edema)，脑肿胀(Brain swelling)，鼻甲肥大(Turbinate
hypertrophy)，鼻中隔偏曲(Nasal septal deviation)，鼻窦黏膜增厚(Sinus mucosal thickening)，鼻腔积液(Nasal cavity fluid)"
  ],
  "impression": [
    "急性脑梗死(Acute cerebral infarction)，硬膜下血肿(Subdural hematoma)，硬膜下血肿(Subdural hematoma)，脑转移瘤(Brain
metastasis)，颅内动脉瘤(Intracranial aneurysm)，蛛网膜下腔出血(Subarachnoid hemorrhage)，真菌性上颌窦炎(Fungal maxillary
sinusitis)，脑膜瘤(Meningioma)，脱髓鞘病变(Demyelinating lesion)，颅骨凹陷性骨折(Depressed skull fracture)，
脑积水(Hydrocephalus)，脑积液(Intracranial fluid collection)，脑积血(Intracranial blood collection)，脑萎缩(Cerebral atrophy)，
脑动脉硬化(Cerebral arteriosclerosis)，轴索损伤(Axonal injury)，筛窦炎(Ethmoid sinusitis)，蝶窦炎(Sphenoid sinusitis)，
额窦炎(Frontal sinusitis)，中耳乳突炎(Otomastoiditis)，乳突积液(Mastoid effusion)，胆脂瘤(Cholesteatoma)，
脑软化灶(Encephalomalacia)，脑白质疏松(Leukoaraiosis)，蛛网膜囊肿(Arachnoid cyst)，大枕大池(Mega cisterna magna)，
眶脂体疝(Orbital fat herniation)"
  ],
  "emergency_signs": [
    "中线移位(Midline shift)，脑疝(Brain herniation)，脑肿胀(Brain swelling)，脑干受压(Brainstem compression)，
颅内压升高(Increased intracranial pressure)，脑沟消失(Effacement of cerebral sulci)，脑室受压(Ventricular compression)，
脑实质出血(Intraparenchymal hemorrhage)，脑脊液漏(Cerebrospinal fluid leak)，颅内积气(Pneumocephalus)，脑镰下疝(Subfalcine
herniation)，脑室扩大伴受压(Ventricular enlargement with compression)，脑膜脑膨出(Meningoencephalocele)，脑干出血(Brainstem
hemorrhage)，颅内血肿(Intracranial hematoma)，脑挫裂伤(Cerebral contusion and laceration)，颅骨凹陷性骨折(Depressed skull
fracture)，脑脊液外渗(Cerebrospinal fluid extravasation)，脑室扩张(Ventricular dilatation)，松果体区囊肿(Pineal region cyst)"
  ]
}
```

**Supplementary Table 3 | Bilingual keyword lexicon of the FIES framework.** The table lists the 122 prespecified clinical keywords used in the FIES framework, comprising 24 Feature terms, 27 Impression terms, and 71 Emergency Signs terms, together with their English translations. The lexicon was constructed exclusively from the training reports, reviewed and refined by five board-certified radiologists, finalized before model evaluation, and applied unchanged across all evaluation cohorts. Lexical variants and synonymous expressions included in the matching procedure are also provided where applicable.
